# Childhood obesity is linked to putative neuroinflammation in brain white matter, hypothalamus, and striatum

**DOI:** 10.1101/2022.09.01.22279506

**Authors:** Zhaolong Li, Amjad Samara, Mary Katherine Ray, Jerrel Rutlin, Cyrus A. Raji, Joshua S. Shimony, Peng Sun, Sheng-Kwei Song, Tamara Hershey, Sarah A. Eisenstein

**Affiliations:** Department of Psychiatry, Washington University in St. Louis School of Medicine, St. Louis, MO 63110, USA; Department of Psychological and Brain Sciences, Washington University in St. Louis, St. Louis, MO 63130, USA; Department of Neurology, Washington University in St. Louis School of Medicine, St. Louis, MO 63110, USA; Mallinckrodt Institute of Radiology, Washington University in St. Louis School of Medicine, St. Louis, MO 63110, USA

**Keywords:** diffusion MRI, hypothalamus, neuroinflammation, obesity, white matter

## Abstract

Neuroinflammation is both a consequence and driver of overfeeding and weight gain in rodent obesity models. Advances in magnetic resonance imaging (MRI) enable investigations of brain microstructure that suggests neuroinflammation in human obesity. To assess the convergent validity across MRI techniques and extend previous findings, we used diffusion basis spectrum imaging (DBSI) to characterize obesity-associated alterations in brain microstructure in 601 children (age 9-11 years) from the Adolescent Brain Cognitive Development^SM^ Study. Compared to children with normal-weight, greater DBSI restricted fraction (RF), reflecting neuroinflammation-related cellularity, was seen in widespread white matter in children with overweight and obesity. Greater DBSI-RF in hypothalamus, caudate nucleus, putamen, and, in particular, nucleus accumbens, correlated with higher baseline body mass index (BMI) and related anthropometrics. Comparable findings were seen in the striatum with a previously reported restriction spectrum imaging (RSI) model. Gain in waist circumference over one and two years related, at nominal significance, to greater baseline RSI-assessed restricted diffusion in nucleus accumbens and caudate nucleus, and DBSI-RF in hypothalamus, respectively. Here we demonstrate that childhood obesity is associated with microstructural alterations in white matter, hypothalamus, and striatum. Our results also support the reproducibility, across MRI methods, of findings of obesity-related putative neuroinflammation in children.

## 1. Introduction

Childhood obesity is a major growing health issue, affecting over 340 million children worldwide in 2016 (World Health Organization, 2021). It is associated with expensive medical costs (Biener et al., 2020), lower quality of life (Killedar et al., 2020), and elevated risk for health complications including adult obesity, type 2 diabetes, and cardiovascular diseases (Liang et al., 2015; Simmonds et al., 2016). Accumulating evidence also identifies childhood obesity as a risk factor for cognitive dysfunction and Alzheimer’s disease in late-life (Tait et al., 2022). Given the brain’s prominent role in regulating feeding and metabolism, it is essential to understand the relationship between obesity and brain health.

Determining which brain regions and networks might be involved in the development and maintenance of childhood obesity could help identify targets for obesity prevention and intervention, thereby mitigating short and long-term health consequences.

Obesity involves a chronic, low-grade, systemic inflammation affecting multiple organs (Gregor & Hotamisligil, 2011). In rodent models of obesity, high-fat diets induce inflammation in the central nervous system, or “neuroinflammation” (Baufeld et al., 2016; Buckman et al., 2013; De Souza et al., 2005; Décarie-Spain et al., 2018; Valdearcos et al., 2017), which in turn cause memory deficits and anxiodepressive behaviors (Beilharz et al., 2016; Décarie-Spain et al., 2018; Pistell et al., 2010). In humans, post-mortem tissue analyses have revealed associations between obesity and increased gliosis in multiple brain regions, including the hypothalamus, a key regulator of feeding and metabolism (Baufeld et al., 2016; Schur et al., 2015). Aimed at assessing brain health *in vivo*, a number of magnetic resonance imaging (MRI) studies have reported associations between obesity and altered brain structure. In adults, higher body mass index (BMI) and visceral fat are consistently linked to lower cortical thickness and smaller prefrontal and basal ganglia volumes (Fernández-Andújar et al., 2021; Gómez-Apo et al., 2021; Raji et al., 2010; Willette & Kapogiannis, 2015), potentially due to neuronal loss consequent of obesity-related neuroinflammation and/or microangiopathy (Gómez-Apo et al., 2021). These relationships are less clear in children (Willette & Kapogiannis, 2015). Adult obesity has also been associated with compromised white matter integrity, reflected by lower diffusion tensor imaging (DTI)-derived fractional anisotropy (FA) and greater mean diffusivity of water, primarily in frontolimbic tracts and the corpus callosum (Daoust et al., 2021; Kullmann et al., 2015; Verstynen et al., 2012). However, opposite findings of greater white matter DTI-FA in obesity have also been noted (Birdsill et al., 2017; Carbine et al., 2020; Dekkers et al., 2019), and the relationship between obesity and white matter integrity in children remains unknown. Importantly, the standard single-tensor DTI model could be confounded by neuroinflammatory processes such as cellularity and edema (Kullmann et al., 2016; Wang et al., 2011, 2015; Winklewski et al., 2018), which may partially explain the mixed pattern of results.

In recent years, studies using multi-compartment diffusion MRI-based methods, though limited in number, have yielded consistent observations of putative neuroinflammation in feeding-related brain regions in obesity (Rapuano et al., 2020, 2022; Samara et al., 2020, 2021). The data-driven multi-tensor diffusion basis spectrum imaging (DBSI) technique models diffusion-weighted signals as a linear combination of discrete anisotropic tensors and isotropic diffusion spectra, enabling the *in vivo* assessment of brain microstructure (Cross & Song, 2017; Wang et al., 2011, 2015). DBSI metrics, though indirectly reflecting true anatomy, have been histopathologically validated as neuroinflammation-sensitive using rodent and human neural tissue in multiple sclerosis (Chiang et al., 2014; Wang et al., 2014; Wang et al., 2011, 2015), epilepsy (Zhan et al., 2018), and optic neuritis (Lin et al., 2017; Yang et al., 2021). Notably, applying DBSI to adults with obesity, we previously observed microstructural alterations in striatal and limbic regions that suggest cellularity, vasogenic edema, and lower apparent axonal and dendritic densities (Samara et al., 2020, 2021), in line with the obesity-related neuroinflammatory phenotype seen in animal and post-mortem human brain studies. In white matter tracts, we found evidence of increased and widespread DBSI-assessed putative neuroinflammation in young and middle-aged adults with obesity across two independent samples (Samara et al., 2020). DBSI has not yet been used to characterize brain microstructure in childhood obesity. However, Rapuano et al., (2020) used restriction spectrum imaging (RSI), which in contrast to DBSI, models isotropic water diffusion components based on the ratio of radial and axial diffusivities (Palmer et al., 2022; White et al., 2013), and observed associations between greater purported striatal cellular density and higher baseline and future waist circumference and BMI in children in the Adolescent Brain Cognitive Development^SM^ (ABCD) Study (Rapuano et al., 2020, 2022). Furthermore, using a non-diffusion method, namely quantitative T2-weighted MRI, studies have reported that longer hypothalamic T2 relaxation time and greater T2 signal intensity, both suggestive of reactive microglial and astrocytic gliosis, relate to higher BMI in adults (Schur et al., 2015; Thaler et al., 2012) and children, including a subset from the ABCD Study^®^ (Sewaybricker et al., 2019; Sewaybricker, Kee, et al., 2021; Sewaybricker, Melhorn, et al., 2021). Convergent findings amongst MRI methods in the same group of children would support the feasibility and reliability of these techniques to assess putative neuroinflammation in childhood obesity.

In this study, we used the baseline ABCD Study^®^ data from 601 children aged 9-11 years (see **section 2.1** for details on sample selection) to test the *a priori* hypotheses that 1) obesity-associated microstructural alterations, including greater putative neuroinflammation-related cellularity (reflected by greater DBSI restricted fraction (RF)) and lower axonal and dendritic densities (reflected by lower DBSI fiber fraction (FF)) that we had observed in white matter and striatum in adults, and in one novel region not yet assessed using diffusion MRI, i.e., the hypothalamus, would also be present in children, and that 2) greater hypothalamic and striatal cellularity (DBSI-RF) would relate to greater baseline waist circumference and BMI metrics in children, similar to the RSI cellular density metric, namely restricted normalized isotropic (RSI-RNI). We also explored associations between baseline DBSI and RSI metrics in the hypothalamus and striatum and one and two-year longitudinal changes in anthropometrics. If our results using DBSI are consistent to those in studies that used RSI and quantitative T2-weighted MRI, they will support the use of non-invasive MRI-based methods to characterize obesity-related putative neuroinflammation *in vivo* in humans, in the absence of histopathological validation.

## 2. Materials and methods

### 2.1. Participants

Participants were from the ABCD Study^®^, a ten-year, 21-site study tracking brain development in a diverse cohort of U.S. children and adolescents (Casey et al., 2018; Garavan et al., 2018; Jernigan et al., 2018). Participants receive annual physical, sociocultural, and behavioral assessments, as well as neuroimaging and bioassays every two years. Institutional review boards at study sites approved study procedures; parents/caregivers provided written consent and children gave verbal assent. The ABCD Study^®^ 2.0.1 release included data from 11,875 participants at baseline and 4,951 participants at one-year follow-up. In addition to the ABCD Study^®^ inclusion/exclusion criteria (Garavan et al., 2018), we excluded participants with 1) missing anthropometric or demographic data at baseline or one-year follow-up; 2) current or past diagnosis of neurological (including cerebral palsy, brain tumor, stroke, aneurysm, brain hemorrhage, intellectual disability, lead poisoning, muscular dystrophy, multiple sclerosis, and others) and psychiatric (including schizophrenia, autism spectrum disorder, attention-deficit hyperactivity disorder, and others) conditions and diabetes, similar to Rapuano et al., (2020); and 3) T1 or diffusion-weighted images (DWIs) that did not pass quality control or had clinically significant incidental findings (Hagler et al., 2019; Li et al., 2021). Also consistent with Rapuano et al., (2020), in order to maximize harmonization of MRI data across sites, only scans performed on Siemens 3T Prisma platforms (Siemens Healthineers AG, Erlangen, Germany) were included. As the ABCD Study^®^ 4.0 release became available during our study, we further included participants with complete data at two-year follow-up to extend exploratory longitudinal analyses. Lastly, because head motion during MRI scans is known to interfere with diffusion tensor model estimation and give spurious correlations (Ling et al., 2012; Yendiki et al., 2014), we excluded participants with excessive head motion (defined as mean DWI framewise displacement ≥ 2.5 mm) and covaried for mean head motion in statistical analyses.

Our inclusion/exclusion criteria selected a total of 1,613 qualifying participants (see **Supplementary Fig. 1** for flowchart). Age and sex-adjusted BMI percentiles at baseline were used to classify participants by weight status (Kuczmarski et al., 2002), including 63 with underweight (BMI < 5^th^ percentile), 1,140 with normal-weight (NW; 5^th^ to < 85^th^ percentiles), 194 with overweight (OW; 85^th^ to < 95^th^ percentiles), and 216 with obesity (OB; ≥ 95^th^ percentile). To achieve balanced group sizes as well as reduce computational cost, we randomly selected 216 NW participants (matched to OB group size) stratified by sex, and included all 194 OW and 216 OB participants. After neuroimaging processing, data from 25 participants were excluded due to missing/incomplete T1 or DWI acquisition, missing field maps, mismatch between field map and DWI dimensions, or missing/unclear DWI directions. The final analytical sample therefore included 212 NW, 187 OW, and 202 OB participants, for a total *n* = 601.

Such sample size is similar to those in recent literature and should afford sufficient power to detect obesity-related microstructural alterations (see **Supplementary Methods** for power analysis) (Jiang et al., 2023; Sewaybricker, Kee, et al., 2021).

### 2.2. Obesity-related measures

Participant waist circumference (WC), weight, and height were measured at baseline and one and two-year follow-ups (Barch et al., 2018). Raw BMI was calculated (weight(lbs)/height(in)2 × 703). BMI *z*-scores corrected for age and sex were computed using the 2000 CDC growth charts (Kuczmarski et al., 2002). These different measures were used to address the concern that a single index may be less reflective of true adiposity and/or sensitive to fat gain in children (Cole et al., 2005; Taylor et al., 2000).

### 2.3. Neuroimaging

#### 2.3.1 MRI acquisition

Details on T1 and DWI acquisition and harmonization across sites are published elsewhere (Casey et al., 2018; Hagler et al., 2019). T1-weighted anatomical images were collected as a 3D T1-weighted inversion prepared RF-spoiled gradient echo scan, with voxel resolution = 1 mm^3^ isotropic. Spin echo echo-planar imaging was used to acquire multi-shell DWIs with the following parameters: total acquisition time = 7:31, repetition time = 4100 ms, time to echo = 88 ms, matrix size = 140 × 140 × 81, flip angle = 90°, acceleration factor = 3, and voxel resolution = 1.7 mm^3^ isotropic. DWIs were imaged with 7 b = 0 frames and 96 gradient directions (b’s = 500, 1000, 2000, and 3000 s/mm^2^ with 6, 15, 15, and 60 directions, respectively).

#### 2.3.2. DWI and DBSI processing

DWIs were corrected for susceptibility-induced distortion, eddy currents, and head motion using FMRIB Software Library (FSL) *topup* and *eddy* (Smith et al., 2004). Multi-tensor DBSI maps were estimated using an in-house script as previously described (Wang et al., 2011, 2015). Leveraging the multi-shell DWI data, DBSI characterizes brain tissue microstructure by partitioning the total water diffusion signal within each image voxel into isotropic and anisotropic compartments. DBSI modeling produced maps of anisotropic fiber fraction (DBSI-FF; reflects axonal/dendritic density), isotropic nonrestricted fraction (*f*(D) at apparent diffusion coefficient (ADC) > 0.3 μm^2^/ms; reflects vasogenic edema/tissue disintegration/extracellular water), and isotropic restricted fraction (DBSI-RF; *f*(D) at 0 < ADC ≤ 0.3 μm^2^/ms; reflects intracellular water/inflammation-related cellularity) (Chiang et al., 2014; Sun et al., 2020; Wang et al., 2015). Details on DBSI model specification are provided in **Supplementary Methods**. Notably, DBSI-FF and RF are consistently lower and greater, respectively, in adult obesity (Samara et al., 2020, 2021) and serve as the neuroinflammation-related microstructural assessment in the current study. DBSI maps were registered to T1 space first using *epi_reg* and a non-diffusion-weighted image, then by applying the transformation matrix to individual maps using *applyxfm*.

#### 2.3.3. Tract-based spatial statistics (TBSS)

Voxel-wise analyses of white matter DBSI-FF and RF were performed using TBSS (Smith et al., 2006). The DTI model was fitted to preprocessed DWIs using FSL *dtifit*, and DTI-FA maps were eroded by one voxel with end slices removed. Cleaned DTI-FA images were nonlinearly registered to the T1-weighted image of a randomly selected NW participant, averaged, and assigned a threshold at FA > 0.2 to create a white matter skeleton, onto which the DBSI-FF and RF maps were projected.

#### 2.3.4. Segmentation of the striatum and hypothalamus

The nucleus accumbens, caudate nucleus, and putamen were segmented from T1-weighted images using FSL *FIRST* (Patenaude et al., 2011). The hypothalamus was segmented using a novel, automated algorithm developed with deep convolutional neural networks trained on adult data (Billot et al., 2020). To assess the algorithm’s accuracy in children, we compared automated and manual hypothalamus segmentations in 20 participants (10 NW and 10 OB, randomly selected within each group). Within this group, the automated and manual segmentations had good spatial overlap (mean Dice similarity coefficient = 0.74, *SD* = 0.02, one-tailed *p* < 0.001 against the conventional threshold of 0.7) and yielded highly correlated volumes (*r* = 0.74, *p* < 0.001). Neither spatial overlap nor volumetric correlation between the automated and manual segmentations was different by weight group (NW vs. OB; *p*’s = 0.58 and 0.97). Although the automated segmentations had smaller volumes than manual segmentations (*means* = 747 and 887 mm^3^, *p* < 0.001), such volume reduction primarily excluded voxels near the hypothalamic surface, reducing possible contamination of diffusion signal from neighboring cerebrospinal fluid and vasculature (**Supplementary Fig. 2**). Also, the segmented volumes were consistent with literature values (Neudorfer et al., 2020). Taken together, the automated algorithm reliably produced hypothalamus segmentations comparable to manual segmentation. For each subcortical structure, segmentations were visually inspected for accuracy before statistical analyses, and volume and DBSI-FF and RF metrics were each extracted and combined/averaged between hemispheres.

### 2.4. Statistical analyses

All analyses, except for TBSS, were performed in R version 4.2.1 (R Core Team, 2013).

Differences in participant characteristics across NW, OW, and OB groups were assessed using analysis of variance (ANOVA) or chi-square tests.

#### 2.4.1. White matter

For TBSS, we excluded data from 28 randomly selected siblings, eliminating family dependency confounds. Baseline DBSI-FF and RF in white mater tracts were compared amongst unrelated NW (*n* = 202), OW (*n* = 180), and OB (*n* = 191) participants using voxel-wise TBSS, first by ANOVAs for main effects of group and second by *t*-tests for between-group comparisons. FSL *Randomize* (null distribution built from 10,000 permutations; with recommended threshold-free cluster enhancement (TFCE)) was used for these comparisons with spatial family-wise error (FWE) rate corrected at two-tailed *p* ≤ 0.05) (Winkler et al., 2014). Briefly, the raw statistical image was TFCE-transformed into an output image in which voxel-wise TFCE scores were weighted sums of local clustered signals, such that larger TFCE scores reflected magnitude of cluster-like spatial support greater than a given height (signal intensity) (Li et al., 2017; Smith & Nichols, 2009). We specified the –T2 option in *Randomize* (2D optimization for skeletonized data, cluster height weighted by *H* = 2, cluster extent weighted by *E* = 1, voxel connectivity = 26). Voxel-wise analyses using TBSS and TFCE allowed for sensitive detection of regionally-specific obesity-related DBSI-FF and RF effects in white matter, while stringently controlling for multiple comparisons across space. Participant age, sex, race/ethnicity, parental education, household income, parental marital status, pubertal development stage (PDS), mean head motion, and intracranial volume (ICV) were covaried in TBSS. Group differences in white matter skeleton-average values of DBSI-FF and RF were assessed with linear mixed-effects models using the *lme4* package (Bates et al., 2015), where the same set of covariates plus weight group were fixed effects and site was the random effect.

#### 2.4.2. Striatum and hypothalamus

DBSI-FF and RF outliers in the nucleus accumbens, caudate nucleus, putamen, and hypothalamus that were ± 3 SD away from the mean were removed (**Supplementary Table 1**). One and two-year changes in obesity-related measures (i.e., WC, BMI, and BMI *z*-scores) were calculated by subtracting baseline from respective follow-up. Extreme BMI values (< 10 kg/m^2^ or > 50 kg/m^2^) and associated BMI *z*-scores were removed, including 1 NW and 1 OB at one-year and 1 OW at two-year. Distributions for obesity-related measures at and changes between all timepoints are shown in **Supplementary Fig. 3**.

Associations between DBSI metrics and baseline or future change in obesity-related measures were assessed using linear mixed-effects models. Age (at baseline, one, or two-year), sex, race/ethnicity, PDS (at baseline, one, or two-year), parental education, household income, parental marital status, mean head motion, and ICV were covaried due to potential confounding (Lawrence et al., 2022; Li et al., 2023; Palmer et al., 2022; Rapuano et al., 2020), and the random effect was family nested within sites. In longitudinal models, baseline obesity-related measures were also covaried. As we had *a priori* hypotheses, and the goal was to describe regionally-specific relations between tissue microstructure and convergent obesity-related measures, multiple comparisons were corrected with each structure treated as a family, at two-tailed *p* = 0.05 / (4 regions × 2 DBSI metrics) = 0.00625. Effect size estimates were standardized *β*’s with 95% confidence intervals (CIs) and partial *R*^2^’s. Models were checked for normality of residuals, homoscedasticity, and low multicollinearity (variance inflation factors were ≤ 2.56). As there were missing data following outlier removal, sample sizes varied and are reported in individual analyses.

#### 2.4.3. Comparison between DBSI and RSI

Mean RSI restricted normalized isotropic (RSI-RNI) metrics in bilateral nucleus accumbens, caudate nucleus, and putamen were obtained from the ABCD Study^®^ tabulated dataset (Hagler et al., 2019). The ABCD Study^®^ segmented structures using FreeSurfer v5.3; the hypothalamus was not specifically segmented and voxel-wise RSI-RNI maps were not available. RSI reflects cellularity as an increase in the restricted isotropic (originating from intracellular water) diffusion signal, i.e., RSI-RNI (Rapuano et al., 2020, 2022). Associations between RSI-RNI and baseline or future change in obesity-related measures were evaluated using linear mixed-effects models, as in DBSI described in **section 2.4.2**. To further compare model performance, DBSI-RF and RSI-RNI from the nucleus accumbens, caudate nucleus, and putamen were each tested on classifying NW and OB participants using mixed-effects logistic regression, with the same fixed and random effect covariates in linear models. Receiver operating characteristic curves and areas-under-the-curve (AUCs) with 95% CIs were computed using the *pROC* package, and AUCs from DBSI-RF and RSI-RNI were compared using DeLong’s test (Robin et al., 2011).

## 3. Results

### 3.1. Sample characteristics

Participant demographics, neuroimaging metrics, and obesity-related measures are described in **Table 1**. Qualitatively, the OW and/or OB groups compared to the NW group had more non-White participants, more advanced pubertal development, lower parental education, household income, and proportion of married parents, higher baseline obesity-related measures, and greater one and two-year gain in WC but decrease in BMI *z*-scores. Groups did not differ significantly in striatal or hypothalamic volumes; these volumes were thus not covaried in addition to ICV in analyses.

**Table 1.** Participant demographics, brain volumes, and obesity-related measures. Statistics are shown as mean ± standard deviation for continuous variables and count (frequency) for categorical data. Variables were assessed at baseline unless otherwise noted. Comparisons were performed using one-way analysis of variance or chi-squared tests as appropriate. The “Other” category under race/ethnicity included participants who were parent/caregiver-identified as American Indian, Alaskan Native, Native Hawaiian, other Pacific Islander, mixed, or otherwise not listed. Abbreviations: NW, children with normal-weight; OW, with overweight; OB, with obesity; PDS, pubertal development stage; HS, high school; GED, General Educational Development; ICV, intracranial volume; V, volume; BMI, body mass index; WC, waist circumference. *, *p* ≤ 0.05; **, *p* ≤ 0.01; ***, *p* ≤ 0.001.

| Variable | Group (total $n = 601$) | | | $p$-value |
| --- | --- | --- | --- | --- |
|  | NW | OW | OB |  |
| $n$ | 212 (35.3% of all) | 187 (31.1% of all) | 202 (33.6% of all) | N/A |
| Age (months) | 121 ± 8 | 121 ± 7 | 120 ± 7 | 0.89 |
| Sex |  |  |  | 0.31 |
| Male | 113 (53.3%) | 98 (52.4%) | 120 (59.4%) |  |
| Female | 99 (46.7%) | 89 (47.6%) | 82 (40.6%) |  |
| Race/ethnicity |  |  |  | < 0.001 *** |
| Asian | 1 (0.5%) | 1 (0.5%) | 4 (2.0%) |  |
| Black | 16 (7.5%) | 23 (12.3%) | 27 (13.4%) |  |
| Hispanic | 25 (11.8%) | 37 (19.8%) | 54 (26.7%) |  |
| White | 156 (73.6%) | 112 (59.9%) | 99 (49.0%) |  |
| Other | 14 (6.6%) | 14 (7.5%) | 18 (8.9%) |  |
| PDS category |  |  |  | < 0.001 *** |
| 1 | 129 (60.8%) | 76 (40.6%) | 86 (42.6%) |  |
| 2 | 53 (25.0%) | 49 (26.2%) | 51 (25.2%) |  |
| 3+ | 30 (14.2%) | 62 (33.2%) | 65 (32.2%) |  |
| Parental education |  |  |  | < 0.001 ** |
| No HS diploma | 0 (0%) | 1 (0.5%) | 4 (2.0%) |  |
| HS diploma/ GED | 7 (3.3%) | 5 (2.7%) | 20 (9.9%) |  |
| Some college | 14 (6.6%) | 28 (15.0%) | 42 (20.8%) |  |
| Bachelor | 97 (45.8%) | 87 (46.5%) | 84 (41.6%) |  |
| Postgraduate | 94 (44.3%) | 66 (35.3%) | 52 (25.7%) |  |
| Household income |  |  |  | < 0.001 *** |
| < 50k | 32 (15.1%) | 45 (24.1%) | 62 (30.7%) |  |
| ≥ 50k & < 100k | 64 (30.2%) | 74 (39.6%) | 74 (36.6%) | |
| ≥ 100k | 116 (54.7%) | 68 (36.4%) | 66 (32.7%) | |
| Parental marriage |  |  |  | 0.005 ** |
| Married | 164 (77.4%) | 128 (68.4%) | 127 (62.9%) |  |
| Not married | 48 (22.6%) | 59 (31.6%) | 75 (37.1%) |  |
| Mean motion (mm) | 1.23 ± 0.25 | 1.27 ± 0.31 | 1.27 ± 0.30 | 0.25 |
| ICV (mm<sup>3</sup>) | 1539012 ± 128794 | 1569834 ± 151931 | 1559140 ± 144992 | 0.09 |
| V<sub>hypothalamus</sub> (mm<sup>3</sup>) | 737 ± 90 | 740 ± 104 | 733 ± 108 | 0.77 |
| V<sub>nucleus accumbens</sub> (mm<sup>3</sup>) | 1028 ± 204 | 1057 ± 225 | 1034 ± 204 | 0.36 |
| V<sub>caudate nucleus</sub> (mm<sup>3</sup>) | 7902 ± 990 | 8038 ± 975 | 7853 ± 998 | 0.16 |
| V<sub>putamen</sub> (mm<sup>3</sup>) | 10609 ± 1154 | 10691 ± 1162 | 10627 ± 1206 | 0.77 |
| Obesity-related measures |  |  |  |  |
| Baseline |  |  |  |  |
| BMI (kg/m<sup>2</sup>) | 16.82 ± 1.39 | 20.92 ± 1.06 | 25.85 ± 3.41 | < 0.001 *** |
| BMI percentile | 48.51 ± 22.38 | 90.43 ± 2.90 | 97.63 ± 1.32 | < 0.001 *** |
| BMI z-score | -0.06 ± 0.65 | 1.33 ± 0.17 | 2.05 ± 0.29 | < 0.001 *** |
| WC (inch) | 25.13 ± 2.27 | 29.16 ± 2.68 | 32.96 ± 4.10 | < 0.001 *** |
| One-year |  |  |  |  |
| BMI (kg/m<sup>2</sup>) | 17.61 ± 1.88 | 21.97 ± 2.24 | 27.16 ± 4.63 | < 0.001 *** |
| BMI z-score | -0.01 ± 0.75 | 1.28 ± 0.47 | 1.98 ± 0.53 | < 0.001 *** |
| WC (in) | 25.93 ± 2.68 | 30.03 ± 2.82 | 34.89 ± 4.48 | < 0.001 *** |
| Change ($\Delta$) over one year | | | | |
| ΔBMI (kg/m<sup>2</sup>) | 0.79 ± 1.29 | 1.05 ± 1.97 | 1.30 ± 3.57 | 0.12 |
| ΔBMI z-score | 0.04 ± 0.51 | -0.05 ± 0.44 | -0.08 ± 0.45 | 0.024 * |
| ΔWC (in) | 0.80 ± 2.34 | 0.88 ± 2.66 | 1.94 ± 3.16 | < 0.001 *** |
| Two-year |  |  |  |  |
| BMI (kg/m<sup>2</sup>) | 18.43 ± 2.32 | 22.91 ± 2.48 | 28.11 ± 4.81 | < 0.001 *** |
| BMI z-score | 0.01 ± 0.86 | 1.28 ± 0.50 | 1.94 ± 0.54 | < 0.001 *** |
| WC (in) | 27.12 ± 3.01 | 31.34 ± 3.27 | 36.02 ± 4.98 | < 0.001 *** |
| Change ($\Delta$) over two years | | | | |
| ΔBMI (kg/m<sup>2</sup>) | 1.62 ± 1.83 | 1.98 ± 2.27 | 2.25 ± 3.85 | 0.07 |
| ΔBMI z-score | 0.07 ± 0.65 | -0.05 ± 0.48 | -0.11 ± 0.48 | 0.002 ** |
| ΔWC (in) | 1.99 ± 2.72 | 2.18 ± 3.01 | 3.06 ± 3.72 | 0.001 ** |

### 3.2. Comparison of white matter DBSI metrics across groups

TBSS ANOVAs indicated significant main effects of weight group for both DBSI-FF and RF. Follow-up TBSS *t*-tests showed that relative to NW, both OW and OB participants had significantly lower DBSI-FF (reflecting lower axonal/dendritic density) and greater DBSI-RF (reflecting elevated cellularity) in widespread white matter tracts (all FWE-corrected *p* ≤ 0.05). Qualitatively, group differences in both DBSI-FF and RF appeared more widespread throughout white matter tracts in the OB vs. NW comparisons than in OW vs. NW comparisons (**Fig. 1**); nonetheless, white matter voxel-wise DBSI-FF and RF were not significantly different between OB and OW groups (FWE-corrected *p* > 0.054). Consistent with voxel-wise comparisons, relative to NW, both OW and OB groups had lower white matter average DBSI-FF (OW vs. NW, *β* = -0.39, 95% CI: -0.60 to -0.18, *p* < 0.001; OB vs. NW, *β* = -0.33, 95% CI: -0.54 to -0.11, *p* = 0.003) and greater DBSI-RF (OW vs. NW, *β* = 0.50, 95% CI: 0.30 to 0.70, *p* < 0.001; OB vs. NW, *β* = 0.36, 95% CI: 0.16 to 0.56, *p* < 0.001), but these differences were not significant between OW and OB (*p*’s = 0.65 and 0.14 for DBSI-FF and RF comparisons).

**Fig. 1.**
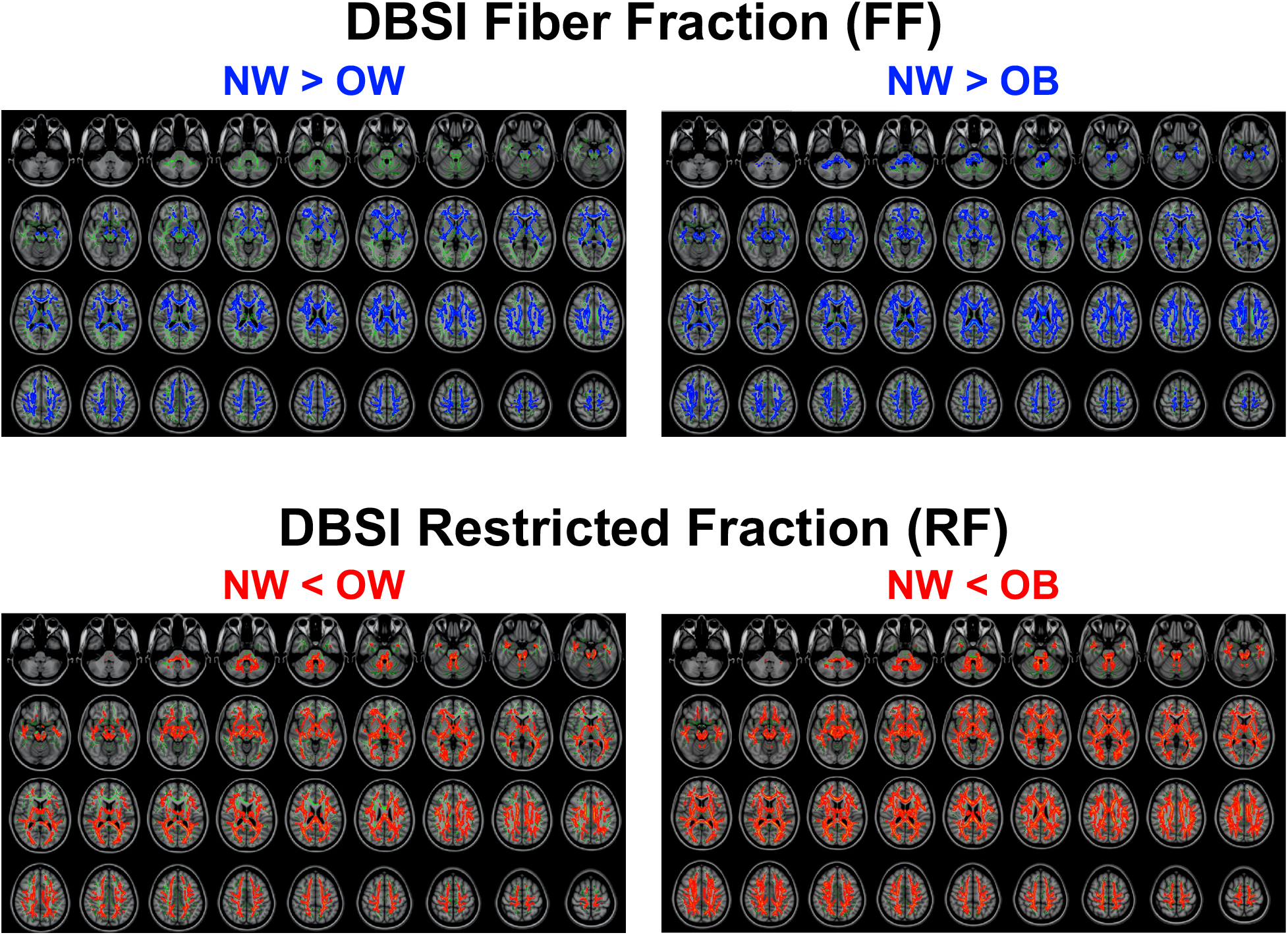
Voxel-wise comparisons of white matter DBSI metrics amongst unrelated children with normal-weight (NW; *n* = 202), overweight (OW, *n* = 180) and obesity (OB; *n* = 191). In each panel, axial images are shown from inferior (top left) to superior (bottom right). In *green*: white matter skeleton; in *blue, light blue*: NW > OB/OW at family-wise error (FWE) rate-corrected *p* ≤ 0.05 and 0.01; in *red, yellow*: NW < OB/OW at FWE-corrected *p* ≤ 0.05 and 0.01. Comparisons were adjusted for age, sex, race/ethnicity, parental education, household income, parental marital status, pubertal development stage, mean head motion, and intracranial volume. DBSI, diffusion basis spectrum imaging.

### 3.3. Associations between striatal and hypothalamic DBSI metrics and obesity-related measures

#### 3.3.1. Baseline

Greater BMI at baseline was significantly associated with greater DBSI-RF in the hypothalamus (*β* = 0.11, 95% CI: 0.04 to 0.19, partial *R*^*2*^ = 0.014, *p* = 0.005), nucleus accumbens (*β* = 0.30, 95% CI: 0.22 to 0.39, partial *R*^*2*^ = 0.096, *p* < 0.001), caudate nucleus (*β* = 0.18, 95% CI: 0.08 to 0.27, partial *R*^*2*^ = 0.034, *p* < 0.001), and the putamen (*β* = 0.14, 95% CI: 0.06 to 0.22, partial *R*^*2*^ = 0.021, *p* = 0.001) (**Fig. 2A**). These results were consistent with WC and BMI *z*-scores as obesity-related measures. Further, greater baseline BMI *z*-scores were significantly related to lower DBSI-FF in the hypothalamus (*β* = - 0.12, 95% CI: -0.20 to -0.05, partial *R*^*2*^ = 0.016, *p* = 0.002) (**Fig. 2B**). Similar associations, at nominal but not multiple comparison-adjusted significance, were seen between lower DBSI-FF in the hypothalamus and WC (*β* = -0.08, *p* = 0.038) and BMI (*β* = -0.09, *p* = 0.024); in the nucleus accumbens and WC (*β* = - 0.10, *p* = 0.009); and in the putamen and BMI (*β* = -0.08, *p* = 0.048) and BMI *z*-scores (*β* = -0.08, *p* = 0.048). Detailed statistics for all models are reported in **Supplementary Table 2**. Follow-up analyses revealed no DBSI metric by hemisphere interaction in relating to baseline obesity-related measures (i.e., no laterality effect; *p*’s ≥ 0.30).

**Fig. 2.**
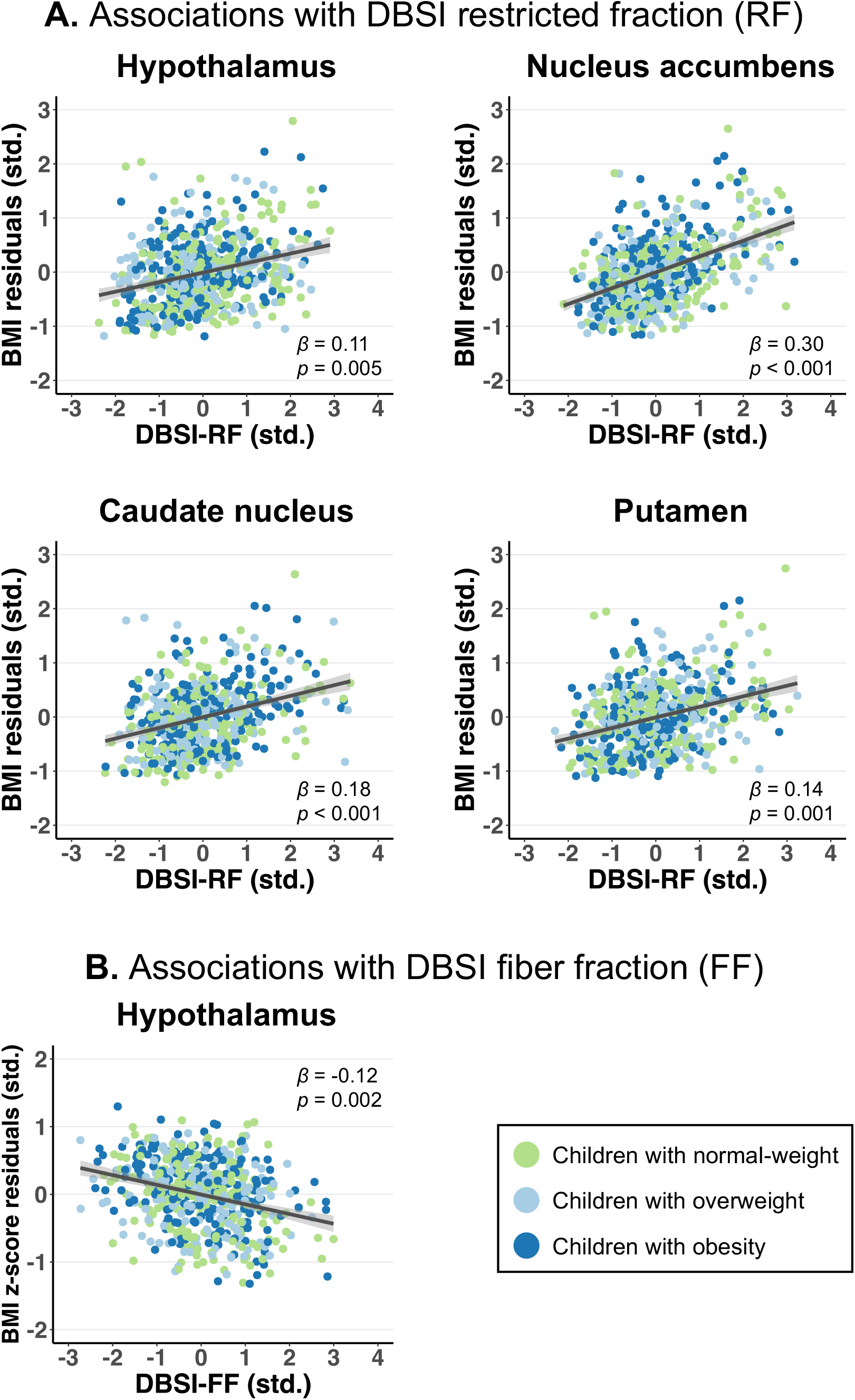
Significant associations **(A)** between baseline body mass index (BMI) and DBSI-RF in the hypothalamus and striatum and **(B)** between baseline BMI *z*-scores and DBSI-FF in the hypothalamus in children. BMI or BMI *z*-score residuals (adjusted for age, sex, race/ethnicity, parental education, household income, parental marital status, pubertal development stage, mean head motion, intracranial volume, and family nested by site) and DBSI metrics were standardized (std.). Standardized *β* regression coefficients were reported with 95% confidence intervals (shaded). DBSI, diffusion basis spectrum imaging.

Beyond DBSI metrics, variables that were associated with greater obesity-related measures at baseline included older age, lower parental education, and more advanced pubertal stage. As we did not specifically power or hypothesize for demographics-related effects, these findings are exploratory and are noted in **Supplementary Table 3**. In total, our linear-mixed effects models explained 18-25% of the variance in baseline obesity-related measures.

#### 3.3.2. One and two-year change

Greater DBSI-RF in the hypothalamus at baseline, at nominal significance not surviving multiple comparison correction, predicted greater gain in WC over two years, accounting for baseline WC (*β* = 0.09, 95% CI: 0.01 to 0.18, partial *R*^*2*^ = 0.008, *p* = 0.035; **Supplementary Fig. 4**). However, such effect was not seen at one-year follow-up, or with changes in BMI or BMI *z*-scores as obesity-related measures (*p*’s = 0.79 and 0.52). Other one or two-year changes in obesity-related measures were not associated with baseline DBSI metrics (**Supplementary Tables 4** and **5**).

### 3.4. Associations between striatal RSI-RNI and obesity-related measures

#### 3.4.1. Baseline

Consistent with a previous study using ABCD Study^®^ baseline data (*n* = 5,366; Rapuano et al., 2020), greater baseline BMI was associated with higher RSI-RNI in the nucleus accumbens (*β* = 0.36, 95% CI: 0.27 to 0.44, partial *R*^*2*^ = 0.125, *p* < 0.001), caudate nucleus (*β* = 0.15, 95% CI: 0.07 to 0.23, partial *R*^*2*^ = 0.025, *p* < 0.001), and putamen (*β* = 0.17, 95% CI: 0.09 to 0.26, partial *R*^*2*^ = 0.030, *p* < 0.001) in the current, smaller sample. Results were similar with WC and BMI *z*-scores. Detailed statistics for all linear mixed-effects models are reported in **Supplementary Table 6**.

#### 3.4.2. One and two-year change

Greater baseline RSI-RNI in the nucleus accumbens and caudate nucleus were respectively associated, not surviving multiple comparison correction, with one-year gain in WC, accounting for baseline levels (nucleus accumbens: *β* = 0.10, 95% CI: 0.00 to 0.20, partial *R*^*2*^ = 0.008, *p* = 0.042; caudate nucleus: *β* = 0.11, 95% CI: 0.02 to 0.19, partial *R*^*2*^ = 0.011, *p* = 0.017; **Supplementary Fig. 5**). These associations were not seen at two-year follow-up or with changes in BMI or BMI *z*-scores (**Supplementary Tables 7** and **8**).

### 3.5. Comparison between DBSI and RSI on classifying NW and OB groups

Striatal DBSI and RSI metrics reflective of neuroinflammation-associated cellularity and cellular density, i.e., DBSI-RF and RSI-RNI, showed similar sensitivity and specificity in classifying NW and OB children (*p*’s ≥ 0.13; **Fig. 3**). Across the striatum, DBSI-RF was positively and strongly correlated with RSI-RNI (*r*’s ≥ 0.60, *p*’s ≤ 0.001; **Supplementary Fig. 6**).

**Fig. 3.**
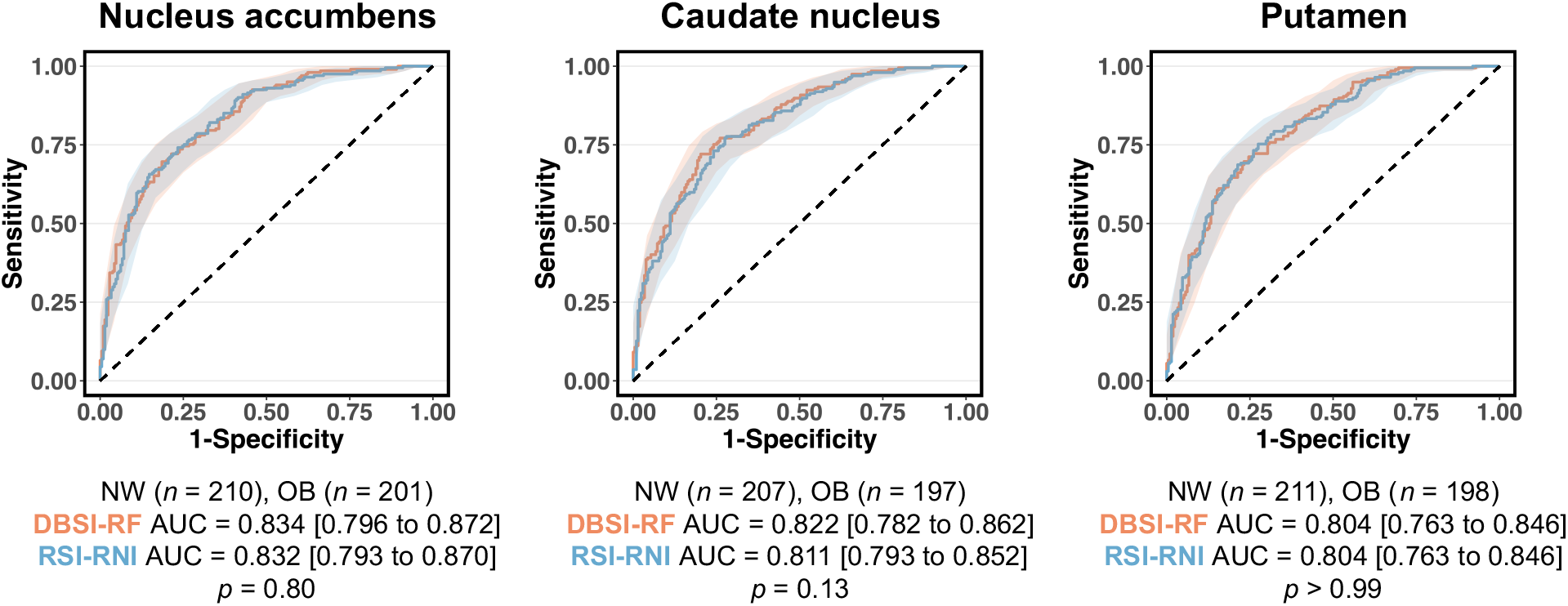
Receiver operating characteristic curves comparing striatal DBSI restricted fraction (RF) and RSI restricted normalized isotropic (RNI) performance in classifying children with normal-weight (NW) and obesity (OB). AUC, area-under-the-curve; DBSI, diffusion basis spectrum imaging; RSI, restriction spectrum imaging.

## 4. Discussion

### 4.1. Overview

Here we present both novel findings and support for the reproducibility of previous neuroimaging studies that observed microstructural alterations suggestive of neuroinflammation in key feeding and reward-related brain regions in childhood obesity. First, we demonstrate that elevated DBSI-assessed cellularity, i.e., putative inflammatory marker, in the striatum relates to higher WC, BMI, and BMI *z*-scores in 601 children aged 9 to 11 years from the ABCD Study^®^, reproducing observations made by Rapuano et al., (2020) that used another diffusion-based RSI model in the same dataset. Quantitatively, DBSI-RF and RSI-RNI were associated with obesity-related measures in similar magnitudes, and the two methods exhibited comparable performance in classifying NW vs. OB children. Such convergence of findings underpins the sensitivity and utility of diffusion MRI-based techniques in characterizing brain microstructural alterations in obesity.

Second, we observed associations between obesity and increased purported cellularity consistent with neuroinflammation in brain white matter tracts and hypothalamus, which were not assessed by Rapuano et al., (2020). Our results in the hypothalamus align with reports of putative gliosis in this region, assessed by quantitative T2 MRI, in both childhood and adult obesity (Schur et al., 2015; Sewaybricker et al., 2019; Sewaybricker, Kee, et al., 2021; Sewaybricker, Melhorn, et al., 2021; Thaler et al., 2012). Here, our findings add diffusion MRI-derived evidence of obesity-related putative neuroinflammation in the hypothalamus in children. Furthermore, to our knowledge, our study is the first to investigate and report overweight and obesity-associated white matter microstructural alterations in children, in line with our earlier studies of DBSI-assessed putative white matter neuroinflammation in adults (Samara et al., 2020). Collectively, our findings and those previously reported suggest that young children manifest obesity-related differences in brain microstructure that are consistent with neuroinflammation seen in animal and post-mortem human brain studies (Baufeld et al., 2016; Buckman et al., 2013; De Souza et al., 2005; Décarie-Spain et al., 2018; Schur et al., 2015; Valdearcos et al., 2017). Such brain differences may affect current and future susceptibility for weight gain and its comorbidities including cognitive impairment, type 2 diabetes, and late-life dementia (Liang et al., 2015; Simmonds et al., 2016; Tait et al., 2022).

### 4.2. Links between obesity, neuroinflammation, and brain function

The highly vascularized hypothalamus responds to feeding-related hormones, neuronal signals, and nutrients derived from the bloodstream (Velloso & Schwartz, 2011). As a “metabolic sensor”, the hypothalamus is vulnerable to overfeeding and obesity-related elevations in peripheral pro-inflammatory molecules including cytokines and saturated fatty acids (Jais & Brüning, 2017). Overfeeding also causes the blood-brain barrier to break down, further enabling inflammatory factors to infiltrate brain tissue (Guillemot-Legris et al., 2016; Guillemot-Legris & Muccioli, 2017; Stranahan et al., 2016). Our finding that DBSI-assessed cellularity (DBSI-RF) in the hypothalamus is greater in childhood obesity is consistent with the neuroinflammatory phenotype encompassing the recruitment, proliferation, and activation of astrocytes and microglia (i.e., reactive gliosis) seen in this brain region in rodents fed with high-fat diets (Buckman et al., 2013; De Souza et al., 2005). While such immune response may initially be neuroprotective, chronic gliosis leads to dysregulated neuroinflammatory processes that disrupt hypothalamic metabolic regulation and contribute to overfeeding, leptin and insulin-resistance, and development of obesity (Gómez-Apo et al., 2021; Sochocka et al., 2017; Valdearcos et al., 2017).

Persistent neuroinflammation could also cause axonal damage and loss (Frischer et al., 2009; Kempuraj et al., 2016), which may explain our observed association between obesity and lower DBSI-assessed axonal/dendritic density (DBSI-FF).

The striatum plays a key role in reward processing and appetitive behavior (Stice et al., 2011). Striatal activity, primarily dopamine neurotransmission, is influenced by homeostatic signals from the hypothalamus and by circulating feeding-related hormones, both acting on receptors on midbrain dopaminergic cells (Abizaid et al., 2006; Figlewicz, 2016; Hommel et al., 2006; King et al., 2011).

Altered dopamine neurotransmission has been noted in obesity (Geiger et al., 2009; Wang et al., 2001; Wu et al., 2017). Beyond the hypothalamus, neuroinflammation in the striatum may further contribute to obesogenic behavior. Indeed, our observation of heightened DBSI-assessed cellularity across the striatum in childhood obesity matches the microstructural changes characteristic of diet-induced reactive gliosis in the nucleus accumbens in rodents (Décarie-Spain et al., 2018; Molina et al., 2020). Taken together, MRI-based assessments of hypothalamic and striatal microstructure by us and others consistently suggest putative neuroinflammation in these regions in childhood obesity, in agreement with studies in rodent models and human adults.

Longitudinally, greater DBSI-assessed cellularity in the hypothalamus weakly predicted two-year gain in WC, aligning with a recent T2 MRI-based report of putative hypothalamic gliosis being associated with weight gain in children (Sewaybricker, Kee, et al., 2021). Further, greater RSI-RNI in the nucleus accumbens and caudate nucleus were linked to one-year WC gain, reproducing findings in Rapuano et al., (2020). However, these findings were at nominal but not multiple comparison-corrected significance, and did not generalize across different obesity-related measures or MRI techniques. As our sample size was not intended to power for the weaker longitudinal effects observed in Rapuano et al., (2020), these findings require confirmation in larger studies involving more longitudinal observations as the ABCD Study^®^ continues to release data. Nonetheless, given evidence that striatal neuroinflammation causally contribute to overfeeding in rodents (Décarie-Spain et al., 2018), plus emerging reports that putative nucleus accumbens cellularity may mediate the relationships between eating behavior and obesity in both adults and children (Rapuano et al., 2022; Samara et al., 2021), chronic neuroinflammation should be evaluated as a potential contributing factor to obesity maintenance.

### 4.3. Brain microstructure in childhood vs. adult obesity

Overall, the pattern of our results in children agrees with DBSI-assessed microstructural alterations seen in adult obesity (Ly et al., 2021; Samara et al., 2020, 2021). Obesity-associated decrease in apparent axonal/dendritic density and increase in cellularity have been observed in white matter in both adults and children. However, the pattern of results in the striatum differs by age. For example, greater putative cellularity in the nucleus accumbens is associated with higher BMI and related metrics in children, but such effect is absent in adults (Samara et al., 2021). Interestingly, it has been noted that in adults, higher BMI is associated with smaller nucleus accumbens volumes (Dekkers et al., 2019; García-García et al., 2020), whereas in children, such association is reversed (García-García et al., 2020; Rapuano et al., 2017) or absent, as is in the current study and another analysis of the ABCD Study^®^ data (Adise et al., 2021). It is possible that as early reactive responses to obesity, striatal cellularity and gliosis would manifest as microstructural but not volumetric alterations in children, while chronic neuroinflammation would over time contribute to vasogenic edema and atrophy seen in adults (Dorrance et al., 2014; Sochocka et al., 2017), as in multiple sclerosis (Kamholz & Garbern, 2005). Furthermore, as executive control regions such as the prefrontal cortex mature later relative to the striatum (Spear, 2000), striatal disruptions may lead to a more dysregulated reward system that influences obesogenic behavior more strongly in children than in adults. As the ABCD Study^®^ collects biennial neuroimaging scans in the same participants from childhood through adulthood using harmonized MRI sequences, future research should capitalize on this longitudinal dataset to delineate obesity-related brain microstructural changes over development.

### 4.4. Comparison between DBSI and RSI findings

Although DBSI and RSI differ in their modeling of brain microstructure, their measures of restricted water diffusion have been interpreted similarly such that the isotropic intracellular water fraction (DBSI-RF and RSI-RNI) is thought to ultimately reflect the degree of neuroinflammation-related immune cell infiltration or tissue cellularity (Cross & Song, 2017; Rapuano et al., 2020, 2022; Wang et al., 2011, 2015). Indeed, in our study, DBSI and RSI-assessed striatal cellularity related similarly to obesity-related measures and strongly with each other, and classified obesity status with comparable performance. A true head-to-head comparison of the microstructural properties reflected by DBSI-RF and RSI-RNI would however warrant a controlled phantom or immunohistological gold standard. In general, the agreeing findings from DBSI and RSI highlight that diffusion MRI-based techniques are sensitive to characterizing obesity-associated microstructural alterations in children, adding a novel neuroimaging tool that assesses putative neuroinflammation *in vivo*.

### 4.5. Limitations

Limitations and future directions include, first, the lack of longitudinal timepoints besides one and two-year follow-ups. It is possible that obesity-related neuroinflammation affects clinical and behavioral outcomes on a timescale larger than two years. Second, as the ABCD Study^®^ does not record obesity duration, we could not assess when and to what extent brain microstructural changes occur relative to obesity onset. Further research tracking children moving from normal-weight to obesity would be useful. Third, as we focused on assessing associations between brain microstructure and obesity-related measures, factors such as sex and socioeconomic status (SES) that likely impact child development and complicate said associations, though controlled for in analyses, were not tested. In terms of sex, girls have greater fat mass and more concentrated trunk adiposity than boys, even at similar BMIs (Wisniewski & Chernausek, 2009). Further, though obesity is associated with elevated serum leptin levels in both sexes, such effect is stronger in girls, who also demonstrate increases in leptin during puberty as opposed to decreases in boys (Falorni et al., 1997). In terms of SES, socioeconomic adversity is a known risk factor for childhood obesity (Hemmingsson, 2018; Vazquez & Cubbin, 2020), with physical inactivity, unhealthy diet, and stress as proposed mediating mechanisms (Caprio et al., 2008; Gebremariam et al., 2017; Hemmingsson, 2018; Mekonnen et al., 2020). Studies have also noted that girls from disadvantaged neighborhoods are more susceptible to obesity compared to boys (Kranjac et al., 2021), and that girls and boys experience differential dietary influences and weight expectations from parents and peers (Caprio et al., 2008; Shah et al., 2020). Regarding brain microstructure, recent analyses using the ABCD Study^®^ data have shown that girls demonstrate greater RSI-assessed cell and neurite density in white matter compared to boys (Lawrence et al., 2022), and lower SES interacts with greater BMI in relating to putative white matter neuroinflammation and smaller brain volumes (Adise et al., 2022; Dennis et al., 2022; Li et al., 2023). Collectively, these results suggest that there exist complex associations between sex, sociocultural forces, and brain microstructure, and future research should adopt an integrative framework to investigate how they may individually and interactively shape obesity development. On a related note, we emphasize growing concerns that current practices of MRI acquisition and quality control may inadvertently exclude participants in less accessible rural areas, from lower SES families, and of racial/ethnic minorities (Ricard et al., 2023). The exclusion of neuroimaging data with excessive head motion, in particular, poses a challenge in obesity research, as greater BMI is causally and genetically linked to increased motion (Beyer et al., 2020). It is possible that our findings may not generalize to children of all sociodemographic backgrounds, and confirmation in large samples of marginalized populations is needed.

Finally, we note the limited interpretability of diffusion MRI-derived microstructural metrics. While DBSI assessments have been histopathologically validated as neuroinflammation-sensitive in inflammatory neurological diseases including human and rodent models of multiple sclerosis (Chiang et al., 2014; Wang et al., 2011, 2015), and rodent optic neuritis (Lin et al., 2017; Yang et al., 2021), validation remains ongoing for obesity. Although the cellularity and axonal density effects inferred from DBSI-modeled water diffusivity agree with the neuroinflammatory phenotype seen in animal models and human post-mortem brain of obesity (Baufeld et al., 2016; Buckman et al., 2013; De Souza et al., 2005; Décarie-Spain et al., 2018; Schur et al., 2015; Valdearcos et al., 2017), we recognize that DBSI, as any MRI technique, is an indirect marker of brain microstructure and could reflect neural development that otherwise do not involve neuroinflammation (Palmer et al., 2022). On a related note, it is challenging to determine whether feeding-related regions such as the hypothalamus and striatum are the only ones involved in obesity-related neuroinflammation, since a true control region in which this phenomenon is definitively absent has not been identified. Such limitation invites future research to evaluate microstructure throughout gray matter as well as study potential interactions between gray and white matter alterations in obesity. The confidence in the validity of MRI-based assessments of obesity-related neuroinflammation could be explored with rodent models and/or human studies using positron emission tomography methods for measuring neuroinflammatory indicators (e.g., astrocyte and microglia activation).

## 5. Conclusions

With DBSI, we observed microstructural alterations in white matter, hypothalamus, and striatum in children with overweight and obesity. Agreement between DBSI and RSI suggested that diffusion MRI is a sensitive and useful tool for assessing obesity-related putative cellularity in children. Given that childhood and adolescence involve substantial brain development, further longitudinal work is warranted to elucidate how early changes in brain microstructure may contribute to obesity and its comorbidities in the long run.

## Supporting information

All supplementary materials

## Data Availability

The ABCD Study data are publicly available through the National Institute of Mental Health Data Archive (https://nda.nih.gov/abcd). The ABCD Study data used in this report came from the ABCD Study Data Release 2.0.1 (DOI: DOI 10.15154/1506087, July 2019) and 4.0 (DOI: 10.15154/1523041, October 2021). The p-code of script used to generate DBSI maps in this study are available upon request, and the developers of DBSI are in the process of publishing an open-source version of the scripts.

## Abbreviations

ABCD^®^: Adolescent Brain Cognitive Development^SM^
ADC: apparent diffusion coefficient
ANOVA: analysis of variance
AUC: area-under-the-curve
BMI: body mass index
CI: confidence interval
DBSI: diffusion basis spectrum imaging
DTI: diffusion tensor imaging
DWI: diffusion-weighted image
FA: fractional anisotropy
FF: fiber fraction
FSL: FMRIB Software Library
FWE: family-wise error
ICV: intracranial volume
MRI: magnetic resonance imaging
NW: children with normal-weight
OB: children with obesity
OW: children with overweight
PDS: pubertal development stage
RF: restricted fraction
RNI: restricted normalized isotropic
RSI: restriction spectrum imaging
SD: standard deviation
SES: socioeconomic status
TBSS: tract-based spatial statistics
TFCE: threshold-free cluster enhancement
WC: waist circumference
WM: white matter

## Author contributions (CRediT)

**Zhaolong Li, BA:** Conceptualization, Data curation, Formal analysis, Methodology, Software, Visualization, Writing – original draft, Writing – review & editing; **Amjad Samara, MD:** Conceptualization, Data curation, Formal analysis, Software, Writing – Review & Editing; **Mary Katherine Ray, PhD:** Conceptualization, Formal analysis, Writing – original draft, Writing – Review & Editing; **Jerrel Rutlin, BS:** Data curation, Software; **Cyrus A. Raji, MD, PhD:** Writing – review & editing; **Joshua S. Shimony, MD, PhD:** Writing – review & editing; **Peng Sun, PhD:** Methodology, Writing – review & editing; **Sheng-Kwei Song, PhD:** Methodology, Writing – review & editing; **Tamara Hershey, PhD:** Conceptualization, Methodology, Supervision, Writing – review & editing; **Sarah A. Eisenstein, PhD:** Conceptualization, Formal analysis, Methodology, Software, Supervision, Validation, Writing – original draft, Writing – review & editing

## Acknowledgements

The authors wish to thank Richard Ni, Jonathan Koller, and Heather Lugar for their assistance with neuroimaging analyses in this study.

## Data availability statement

The ABCD Study^®^ data are publicly available through the National Institute of Mental Health Data Archive (https://nda.nih.gov/abcd). The ABCD Study^®^ data used in this report came from the ABCD Study^®^ Data Release 2.0.1 (DOI: DOI 10.15154/1506087, July 2019) and 4.0 (DOI: 10.15154/1523041, October 2021). The p-code of script used to generate DBSI maps in this study are available upon request, and the developers of DBSI are in the process of publishing an open-source version of the scripts.

