## Supplementary material for "Childhood obesity is linked to putative neuroinflammation in brain white matter, hypothalamus, and striatum": All supplementary materials

Zhaolong Li, BA; Amjad Samara, MD; Mary Katherine Ray, PhD; Jerrel Rutlin, BS; Cyrus A. Raji, MD, PhD; Joshua S. Shimony, MD, PhD; Peng Sun, PhD; Sheng-Kwei Song, PhD; Tamara Hershey, PhD*; Sarah A. Eisenstein, PhD

***Corresponding author:**
Tamara Hershey, PhD
James S. McDonnell Professor of Cognitive Neuroscience
Department of Psychiatry and Mallinckrodt Institute of Radiology
Washington University in St. Louis School of Medicine
St. Louis, MO 63110
MSC 8134-0070-02


**Table of content** (in order of appearance in main text):


| Page | Designation | Description |
|---|---|---|
| 1 | **Supplementary Fig. 1** | Participant selection flowchart |
| 2 | **Supplementary Methods** | Power analysis for detecting obesity-related microstructural alterations; DBSI model specification |
| 3 | **Supplementary Fig. 2** | Assessment of automated hypothalamus segmentation |
| 4 | **Supplementary Table 1** | DBSI-FF and RF outliers in the striatum and hypothalamus |
| 5-6 | **Supplementary Fig. 3** | Distributions of obesity-related measures |
| 7 | **Supplementary Table 2** | Associations between striatal and hypothalamic DBSI metrics and obesity-related measures at baseline |
| 8-9 | **Supplementary Table 3** | Sample exploratory associations between demographics and baseline obesity-related measures |
| 10 | **Supplementary Fig. 4** | Association between baseline DBSI-RF in the hypothalamus and gain in WC over two years |
| 11 | **Supplementary Table 4** | Associations between baseline striatal and hypothalamic DBSI metrics and one-year change (Δ) in obesity-related measures |
| 12 | **Supplementary Table 5** | Associations between baseline striatal and hypothalamic DBSI metrics and two-year change (Δ) in obesity-related measures |
| 13 | **Supplementary Table 6** | Associations between striatal RSI-RNI and obesity-related measures at baseline |
| 14 | **Supplementary Fig. 5** | Association between baseline RSI-RNI in the nucleus accumbens and caudate nucleus and gain in WC over one year |
| 15 | **Supplementary Table 7** | Associations between baseline striatal RSI-RNI and one-year change (Δ) in obesity-related measures |
| 16 | **Supplementary Table 8** | Associations between baseline striatal RSI-RNI and two-year change (Δ) in obesity-related measures |
| 17 | **Supplementary Fig. 6** | Bivariate correlations between DBSI-RF and RSI-RNI in the striatum |

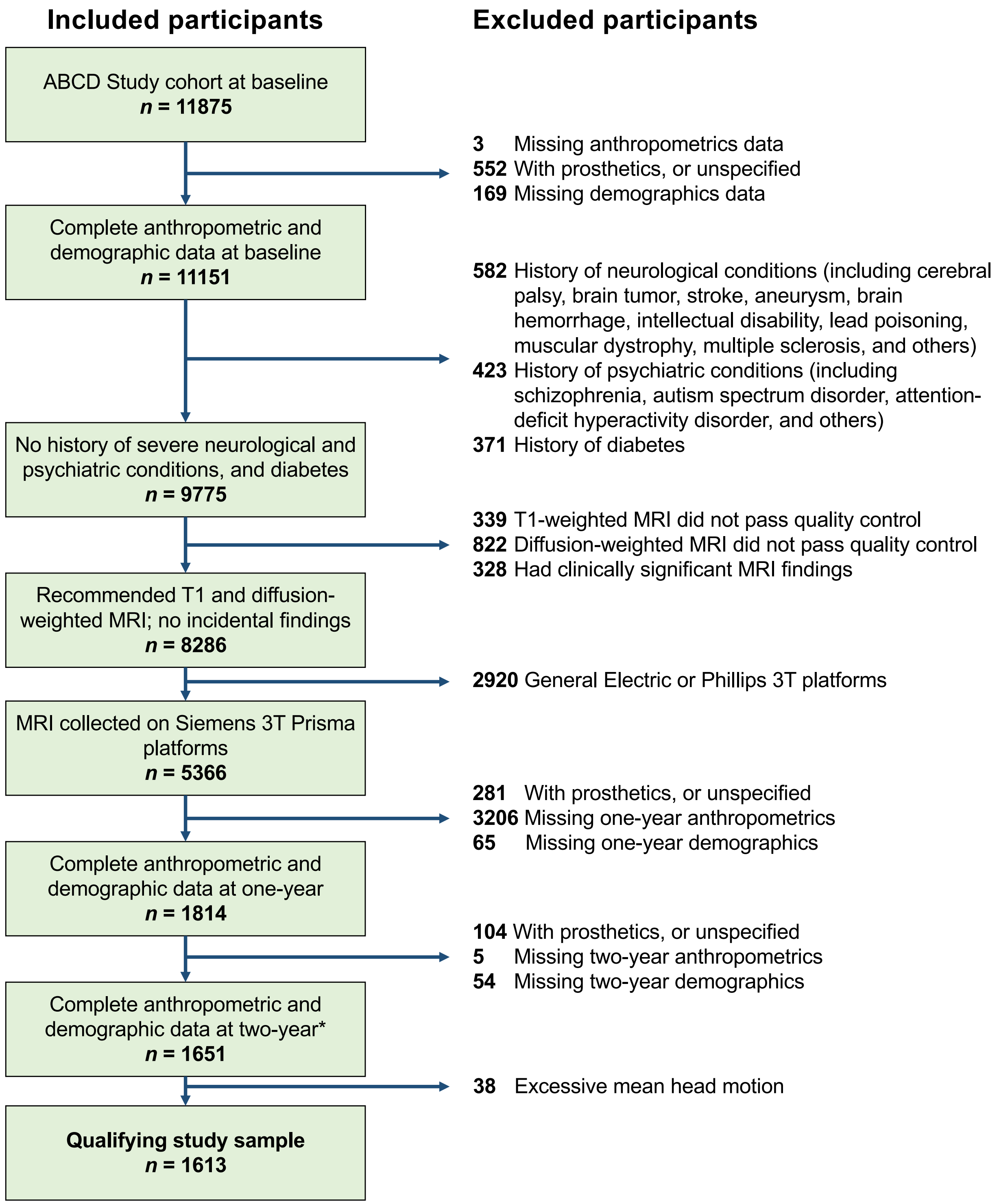


**Supplementary Fig. 1.** Participant selection flowchart. *Our study used data from the ABCD Study® 2.0.1 release, which included assessments for 11,875 participants at baseline and 4,951 participants at one-year follow-up. The inclusion/exclusion criteria here applied to the ABCD Study® 2.0.1 release. However, as the ABCD Study® 4.0 release became available near the end of our initially planned study, we included its two-year data to extend exploratory longitudinal analyses. ABCD, Adolescent Brain Cognitive Development; MRI, magnetic resonance imaging

**Supplementary Methods**

**Power analysis.** Data from 1,613 participants were qualified under our inclusion/exclusion criteria, of which 63 were with underweight, 1,140 were with normal-weight (NW), 194 were with overweight (OW), and 216 were with obesity (OB). To adequately sample for the spectrum of values of obesity-related measures, we processed data for all OW and OB participants, barring those with missing/incomplete/erroneous files. Processing raw DICOM images from the ABCD Study® 2.0.1 release involved time-consuming and computationally heavy steps that included preprocessing and segmenting T1-weighted images, distortion correction and diffusion tensor imaging (DTI) model-fitting for diffusion-weighted images, estimation of diffusion basis spectrum imaging (DBSI) maps, tract-based spatial statistics (TBSS), and quality control, which precluded the processing of all participants in a reasonable timeframe. To streamline analyses, we randomly selected 216 of the qualifying 1,140 NW participants stratified by sex, such that its groups size would be the same to that of the OB group. We reviewed the effect sizes for obesity-related alterations in brain microstructure, including DBSI measurements in white matter tracts and nucleus accumbens in adults (Cohen's *d*'s ≥ 0.7) (Samara et al., 2020, 2021), restriction spectrum imaging (RSI) assessments in striatal regions in children (Cohen's *d*'s ~ 0.5) (Rapuano et al., 2020), and quantitative T2-weighted magnetic resonance imaging (MRI) assessments in the hypothalamus in children and adults (Cohen's *d*'s ≥ 0.65) (Sewaybricker et al., 2019; Sewaybricker, Kee, et al., 2021; Sewaybricker, Melhorn, et al., 2021). A power analysis based on these reported effect sizes indicated that a total sample size of 140 would provide 0.8 statistical power at $\alpha = 0.05$ to detect obesity-related microstructural alterations as assessed by DBSI. Our final analytical sample of $n = 601$ should thus have sufficient power. Further, such sample size is comparable to those in recent literature using the ABCD Study® data (Jiang et al., 2023; Sewaybricker, Kee, et al., 2021).

**Diffusion basis spectrum imaging (DBSI) model specification.** DBSI maps were estimated using in-house scripts as developed in Wang et al., (2011). Compared to traditional single-tensor DTI modeling, DBSI considers the total diffusion signal within each voxel to be a linear combination of multiple anisotropic tensors and a spectrum of isotropic diffusion, thereby resolving their confounding as is common in the imaging of neuroinflammation. The DBSI equation is specified below:

$$S_k = \sum_{i=1}^{N_{Aniso}} f_i e^{-|\overrightarrow{b_k}|.\lambda_{\perp i}} e^{-|\overrightarrow{b_k}|.(\lambda_{\parallel_i} - \lambda_{\perp_i}).\cos^2 \Phi_{ik}} + \int_a^b f(D) e^{-|\overrightarrow{b_k}|D} dD \ (k = 1,2,3, \dots, k)$$

where $S_k$ is the diffusion signal and $b_k$ is the *b*-value of the $k^{th}$ diffusion gradient, $N_{Aniso}$ is the number of anisotropic tensors to be determined, $\Phi_{ik}$ is the angle between the $k^{th}$ diffusion gradient and the principal direction of the $i^{th}$ anisotropic tensor, $\lambda_{\parallel_i}$ and $\lambda_{\perp_i}$ are the axial and radial water diffusivities of the $i^{th}$ anisotropic tensor, $f_i$ is the signal intensity fraction for the $i^{th}$ anisotropic tensor, and *a* and *b* are low and high diffusivity limits of the isotropic diffusion spectrum $f(D)$.

DBSI further partitions the isotropic diffusion spectrum based on *D* cutoffs (i.e., diffusion scales) that empirically corresponded to water diffusion properties in different tissue microstructures, including isotropic nonrestricted fraction ($D > 0.3$ μm$^2$/ms; reflects vasogenic edema/tissue disintegration/extracellular water), and isotropic restricted fraction (DBSI-RF; $0 < D \leq 0.3$ μm$^2$/ms; reflects intracellular water/inflammation-related cellularity) (Wang et al., 2011, 2015). In the current study, DBSI-RF and one anisotropic metric, fiber fraction (DBSI-FF; reflects apparent axonal/dendritic density) served as assessments for brain microstructure. DBSI-FF and RF are consistently lower and higher, respectively, in adults with obesity (Samara et al., 2020, 2021).

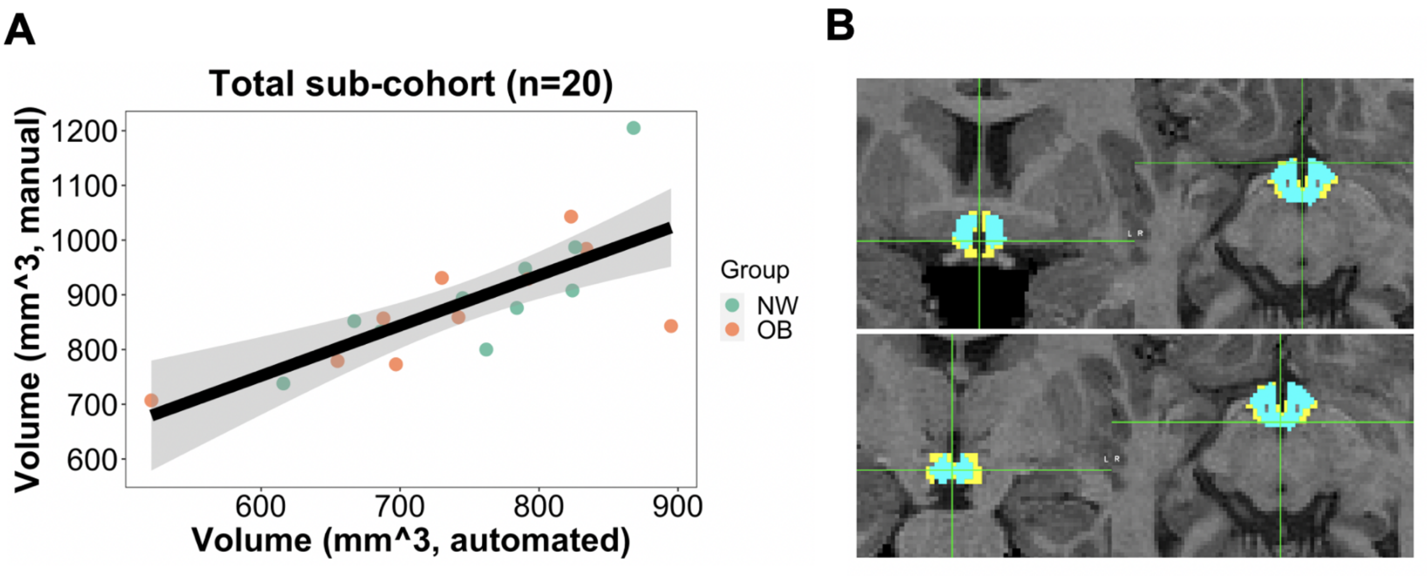


**Supplementary Fig. 2.** Assessment of automated hypothalamus segmentation. **(A)** Volumetric correlation of automated and manual hypothalamus segmentations in 10 randomly selected normal-weight (NW) and 10 randomly selected participants with obesity (OB). **(B)** Sample manual (in yellow) and automated (in blue) segmentations for an OB participant (*first row*, anterior slice in coronal (left) and axial (right) views; *second row*, posterior slice in coronal (left) and axial (right) views).

| Region | Outlier counts | | |
|---|---|---|---|
| | DBSI-FF | DBSI-RF | RSI-RNI |
| Hypothalamus | 6 [1.00%]<br>1 NW, 2 OW, 3 OB | 7 [1.16%]<br>2 NW, 3 OW, 2 OB | N/A |
| Nucleus accumbens | 3 [0.50%]<br>1 OW, 2 OB | 4 [0.67%]<br>2 NW, 1 OW, 1 OB | 2 [0.33%]<br>1 NW, 1 OW |
| Caudate nucleus | 3 [0.50%]<br>1 OW, 2 OB | 9 [1.50%]<br>4 NW, 2 OW, 3 OB | 7 [1.16%]<br>2 NW, 3 OW, 2 OB |
| Putamen | 4 [0.67%]<br>2 OW, 2 OB | 6 [1.00%]<br>1 NW, 1 OW, 4 OB | 1 [0.17%]<br>1 OB |

**Supplementary Table 1. DBSI-FF and RF outliers in the striatum and hypothalamus.** Outliers were defined as ± 3 standard deviations from the mean and were removed in analyses. Tallies are shown as count [frequency], followed by counts by weight groups. RSI-RNI data was not available for the hypothalamus. DBSI, diffusion basis spectrum imaging; FF, fiber fraction; RF, restricted fraction; RSI, restriction spectrum imaging; RNI, restricted normalized isotropic; NW, children with normal-weight; OW, children with overweight; OB, children with obesity.

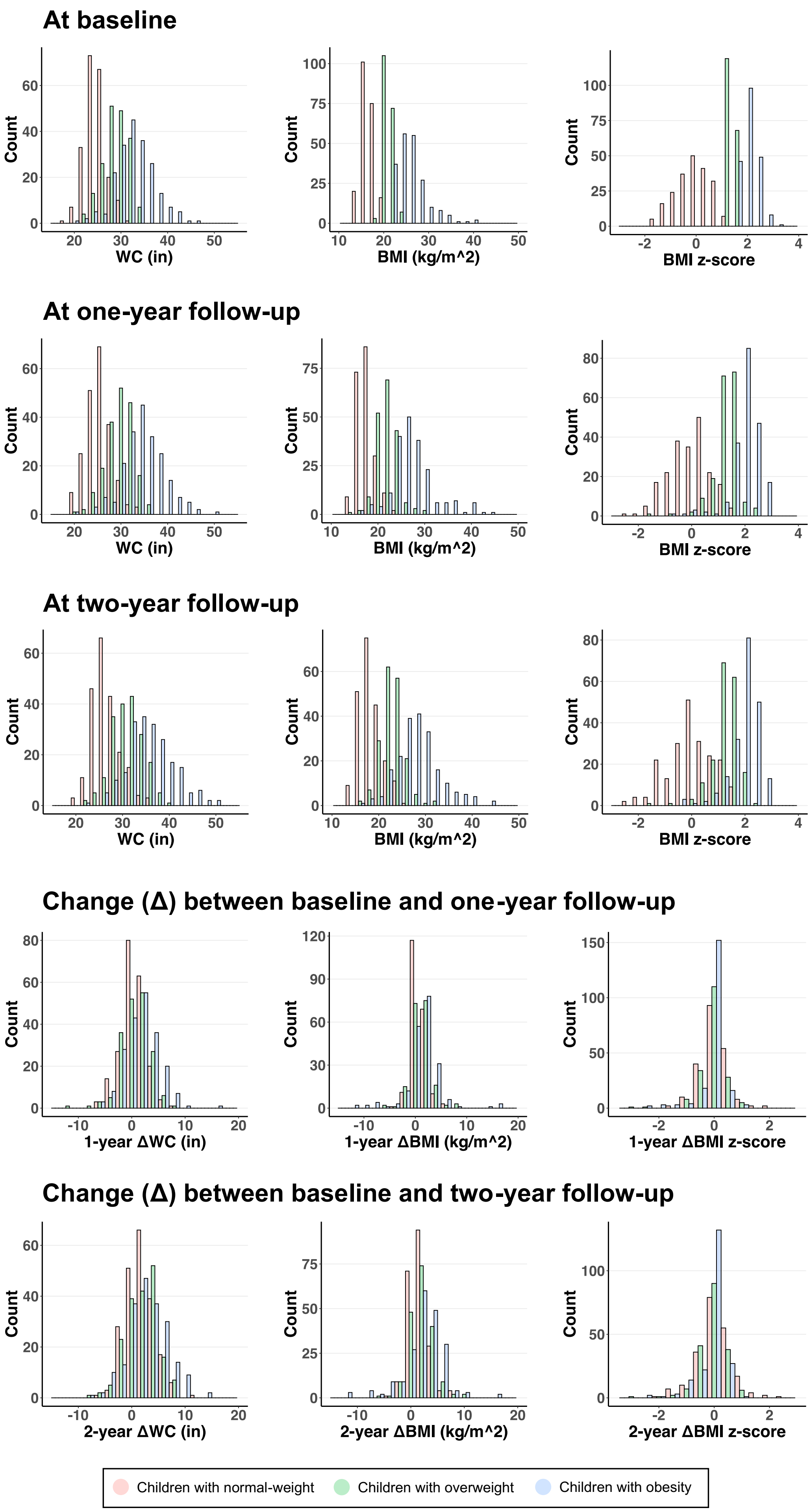
At baseline
Count
WC (in)
BMI (kg/m^2)
BMI z-score
At one-year follow-up
WC (in)
BMI (kg/m^2)
BMI z-score
At two-year follow-up
WC (in)
BMI (kg/m^2)
BMI z-score
Change (Δ) between baseline and one-year follow-up
1-year ΔWC (in)
1-year ΔBMI (kg/m^2)
1-year ΔBMI z-score
Change (Δ) between baseline and two-year follow-up
2-year ΔWC (in)
2-year ΔBMI (kg/m^2)
2-year ΔBMI z-score
Children with normal-weight
Children with overweight
Children with obesity

**Supplementary Fig. 3.** Distributions of obesity-related measures at baseline, one, and two-year follow-ups, and changes (Δ) between timepoints. $N = 601$ for 212 children with normal-weight, 187 with overweight, and 202 with obesity, except that extreme BMI values (< 10 kg/m$^2$ or > 50 kg/m$^2$) and corresponding BMI *z*-scores were removed for 1 participant with normal-weight and 1 participant with obesity at one-year and 1 participant with overweight at two-year. WC, waist circumference (in inches); BMI, body mass index (in kg/m$^2$).

| Region | Imaging metric (predictor) | Baseline obesity-related measure (response) | *p*-value | Std. *β* (95% CI) | Partial $R^2$ | N |
|---|---|---|---|---|---|---|
| **Hypothalamus** | DBSI-FF | WC | 0.038 | -0.08 [-0.15 to -0.01] | 0.007 | 595 |
| | | BMI | 0.024 | -0.09 [-0.16 to -0.01] | 0.009 | 595 |
| | | BMI *z*-score | **0.002** | -0.12 [-0.20 to -0.05] | 0.016 | 595 |
| | DBSI-RF | WC | 0.011 | 0.10 [0.03 to 0.18] | 0.012 | 594 |
| | | BMI | **0.005** | 0.11 [0.04 to 0.19] | 0.014 | 594 |
| | | BMI *z*-score | **0.004** | 0.12 [0.04 to 0.20] | 0.015 | 594 |
| **Nucleus accumbens** | DBSI-FF | WC | 0.009 | -0.10 [-0.17 to -0.03] | 0.011 | 598 |
| | | BMI | 0.09 | -0.07 [-0.14 to 0.01] | 0.005 | 598 |
| | | BMI *z*-score | 0.38 | -0.03 [-0.11 to 0.04] | 0.001 | 598 |
| | DBSI-RF | WC | **< 0.001** | 0.34 [0.25 to 0.42] | 0.113 | 597 |
| | | BMI | **< 0.001** | 0.30 [0.22 to 0.39] | 0.096 | 597 |
| | | BMI *z*-score | **< 0.001** | 0.26 [0.17 to 0.35] | 0.070 | 597 |
| **Caudate nucleus** | DBSI-FF | WC | 0.11 | -0.06 [-0.14 to 0.01] | 0.004 | 598 |
| | | BMI | 0.07 | -0.07 [-0.15 to 0.01] | 0.006 | 598 |
| | | BMI *z*-score | 0.06 | -0.07 [-0.15 to 0.00] | 0.006 | 598 |
| | DBSI-RF | WC | **< 0.001** | 0.23 [0.13 to 0.33] | 0.054 | 592 |
| | | BMI | **< 0.001** | 0.18 [0.08 to 0.27] | 0.034 | 592 |
| | | BMI *z*-score | **0.002** | 0.15 [0.06 to 0.24] | 0.023 | 592 |
| **Putamen** | DBSI-FF | WC | 0.14 | -0.06 [-0.13 to 0.02] | 0.004 | 597 |
| | | BMI | 0.048 | -0.08 [-0.16 to 0.00] | 0.007 | 597 |
| | | BMI *z*-score | 0.048 | -0.08 [-0.16 to 0.00] | 0.007 | 597 |
| | DBSI-RF | WC | **< 0.001** | 0.15 [0.06 to 0.23] | 0.023 | 595 |
| | | BMI | **0.001** | 0.14 [0.06 to 0.22] | 0.021 | 595 |
| | | BMI *z*-score | 0.015 | 0.11 [0.02 to 0.19] | 0.012 | 595 |

**Supplementary Table 2. Associations between striatal and hypothalamic DBSI metrics and obesity-related measures at baseline.** Standardized (Std.) *β* regression coefficients were reported with 95% confidence intervals (CIs). Analyses were adjusted for age, sex, race/ethnicity, parental education, household income, parental marital status, pubertal development stage, mean head motion, intracranial volume, and family nested by site. Effects that survived multiple comparison corrected $p \leq 0.00625$ were considered statistically significant and are bolded. DBSI, diffusion basis spectrum imaging; FF, fiber fraction; RF, restricted fraction; WC, waist circumference; BMI, body mass index.

| (A) Associations between hypothalamus DBSI-RF and baseline BMI (*n* = 594) | | | | |
|---|---|---|---|---|
| **Fixed effects** | **Unstandardized estimate** | **Standard error** | ***p*-value** | **Partial $R^2$** |
| (Intercept) | 6.5e+00 | 3.3e+00 | 0.051 | |
| DBSI-RF | 6.1e+01 | 2.2e+01 | **0.005** | 0.014 |
| Age | 5.1e-02 | 2.3e-02 | **0.025** | 0.009 |
| Male sex | 6.1e-01 | 4.6e-01 | 0.190 | 0.003 |
| Black race/ethnicity (relative to White) | 5.8e-01 | 6.1e-01 | 0.347 | 0.002 |
| Hispanic race/ethnicity (relative to White) | 9.7e-01 | 4.9e-01 | **0.047** | 0.008 |
| Asian race/ethnicity (relative to White) | 4.5e+00 | 1.6e+00 | **0.006** | 0.013 |
| Other race/ethnicity (relative to White) | 8.2e-01 | 6.3e-01 | 0.191 | 0.003 |
| Parental education < HS (relative to Bachelor's) | 1.6e+00 | 1.8e+00 | 0.379 | 0.001 |
| Parental education = HS/GED (relative to Bachelor's) | 1.7e+00 | 8.0e-01 | **0.032** | 0.008 |
| Parental education = Some college (relative to Bachelor's) | 9.6e-01 | 5.2e-01 | 0.064 | 0.006 |
| Parental education = Postgraduate (relative to Bachelor's) | -5.4e-01 | 4.1e-01 | 0.187 | 0.003 |
| Income < 50k (relative to >100k) | 5.3e-01 | 4.1e-01 | 0.204 | 0.001 |
| Income = 50-100k (relative to >100k) | 3.4e-01 | 5.8e-01 | 0.562 | 0.003 |
| Parents not married | 3.7e-01 | 4.3e-01 | 0.391 | 0.001 |
| PDS = 2 (relative to PDS = 1) | 7.8e-01 | 4.1e-01 | 0.057 | 0.006 |
| PDS = 3+ (relative to PDS = 1) | 2.8e+00 | 5.4e-01 | **4e-07** | 0.042 |
| Mean head motion | 9.4e-02 | 5.9e-01 | 0.875 | < 0.001 |
| ICV | 2.6e-06 | 1.3e-06 | 0.055 | 0.006 |
| Total model | | | | 0.179 |
| **(B) Associations between caudate nucleus DBSI-RF and baseline BMI (*n* = 592)** | | | | |
| **Fixed effects** | **Unstandardized estimate** | **Standard error** | ***p*-value** | **Partial $R^2$** |
| (Intercept) | 4.8e+00 | 3.4e+00 | 0.159 | |
| DBSI-RF | 2.0e+02 | 5.2e+01 | **< 0.001** | 0.034 |
| Age | 5.5e-02 | 2.3e-02 | **0.017** | 0.010 |
| Male sex | 8.1e-01 | 4.7e-01 | 0.083 | 0.005 |
| Black race/ethnicity (relative to White) | 1.0e+00 | 6.2e-01 | 0.103 | 0.005 |
| Hispanic race/ethnicity (relative to White) | 8.5e-01 | 4.9e-01 | 0.086 | 0.006 |
| Asian race/ethnicity (relative to White) | 3.9e+00 | 1.6e+00 | **0.016** | 0.010 |
| Other race/ethnicity (relative to White) | 8.5e-01 | 6.3e-01 | 0.179 | 0.003 |
| Parental education < HS (relative to Bachelor's) | 1.9e+00 | 1.8e+00 | 0.304 | 0.002 |
| Parental education = HS/GED (relative to Bachelor's) | 1.8e+00 | 8.1e-01 | **0.030** | 0.008 |
| Parental education = Some college (relative to Bachelor's) | 1.3e+00 | 5.1e-01 | **0.010** | 0.011 |
| Parental education = Postgraduate (relative to Bachelor's) | -4.8e-01 | 4.1e-01 | 0.234 | 0.002 |
| Income < 50k (relative to >100k) | 5.8e-01 | 4.1e-01 | 0.159 | < 0.001 |
| Income = 50-100k (relative to >100k) | 2.1e-01 | 5.8e-01 | 0.714 | 0.003 |
| Parents not married | 2.7e-01 | 4.3e-01 | 0.529 | 0.001 |
| PDS = 2 (relative to PDS = 1) | 7.7e-01 | 4.1e-01 | 0.065 | 0.005 |
| PDS = 3+ (relative to PDS = 1) | 2.8e+00 | 5.4e-01 | **3.78e-07** | 0.042 |

| | | | | |
|---|---|---|---|---|
| Mean head motion | -1.6e-01 | 6.2e-01 | 0.798 | < 0.001 |
| ICV | 2.0e-06 | 1.3e-06 | 0.139 | 0.004 |
| Total model | | | | 0.200 |

**Supplementary Table 3. Sample exploratory associations between demographics and baseline obesity-related measures.** Linear mixed-effects model outputs for associations between baseline body mass index (BMI) and DBSI-RF in the **(A)** hypothalamus and **(B)** caudate nucleus. Similar outputs were seen with BMI *z*-scores and waist circumference, and in other brain regions. Besides DBSI metrics, variables that showed associations with greater obesity-related measures included older age, lower parental education, and more advanced pubertal development stage (PDS). Although there were significant effects of Asian race/ethnicity, its relatively small group size ($n$ = 1, 1, and 4 in normal-weight, overweight, and obesity groups; see **Table 1**) precludes reliable statistical inference. These exploratory results suggest a relationship between demographics and obesity that was controlled for but not tested in the current study. Effects that were nominally significant at $p \leq 0.05$ were bolded. DBSI, diffusion basis spectrum imaging; RF, restricted fraction; HS, high school; GED, General Education Development; ICV, intracranial volume.

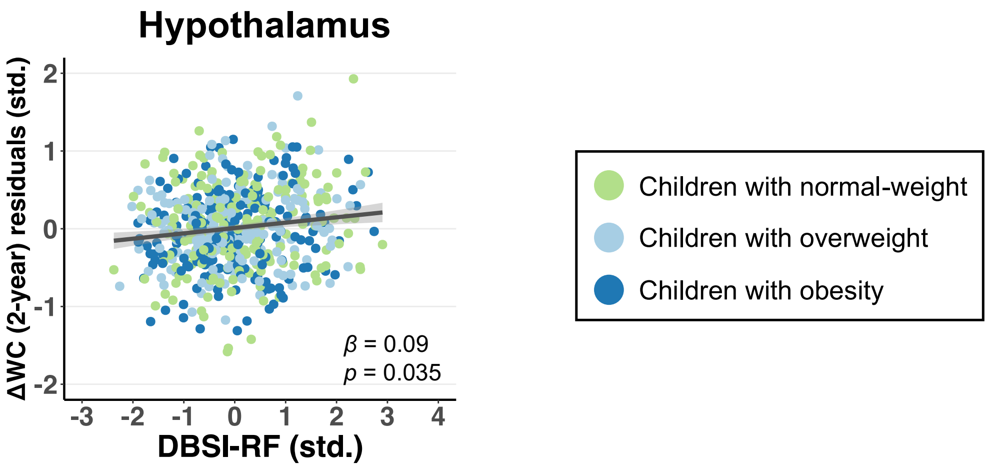


**Supplementary Fig. 4.** Association between baseline DBSI-RF in the hypothalamus and gain in waist circumference (WC) over two years. WC change (Δ) residuals (adjusted for baseline WC, age at two-year, sex, race/ethnicity, parental education, household income, parental marital status, pubertal development stage at two-year, mean head motion, intracranial volume, and family nested by site) and DBSI-RF were standardized (std.). Standardized $\beta$ regression coefficients were reported with 95% confidence intervals (shaded). DBSI, diffusion basis spectrum imaging; RF, restricted fraction.

| Region | Imaging metric (predictor) | 1-year Δobesity-related measure (response) | *p*-value | Std. *β* (95% CI) | Partial $R^2$ | N |
|---|---|---|---|---|---|---|
| **Hypothalamus** | DBSI-FF | WC | 0.63 | -0.02 [-0.10 to 0.06] | < 0.001 | 595 |
| | | BMI | 0.34 | 0.04 [-0.04 to 0.13] | 0.002 | 593 |
| | | BMI *z*-score | 0.89 | 0.01 [-0.08 to 0.09] | < 0.001 | 593 |
| | DBSI-RF | WC | 0.26 | 0.05 [-0.04 to 0.13] | 0.002 | 594 |
| | | BMI | 0.97 | -0.01 [-0.09 to 0.07] | < 0.001 | 592 |
| | | BMI *z*-score | 0.49 | 0.03 [-0.06 to 0.11] | 0.001 | 592 |
| **Nucleus accumbens** | DBSI-FF | WC | 0.89 | -0.01 [-0.09 to 0.08] | < 0.001 | 598 |
| | | BMI | 0.74 | 0.01 [-0.06 to 0.09] | < 0.001 | 596 |
| | | BMI *z*-score | 0.95 | 0.00 [-0.08 to 0.08] | < 0.001 | 596 |
| | DBSI-RF | WC | 0.65 | 0.02 [-0.07 to 0.12] | < 0.001 | 597 |
| | | BMI | 0.88 | 0.01 [-0.09 to 0.08] | < 0.001 | 595 |
| | | BMI *z*-score | 0.85 | 0.01 [-0.08 to 0.09] | < 0.001 | 595 |
| **Caudate nucleus** | DBSI-FF | WC | 0.30 | 0.04 [-0.04 to 0.013] | 0.002 | 598 |
| | | BMI | 0.75 | 0.01 [-0.07 to 0.10] | < 0.001 | 596 |
| | | BMI *z*-score | 0.59 | 0.02 [-0.06 to 0.10] | < 0.001 | 596 |
| | DBSI-RF | WC | 0.47 | 0.04 [-0.07 to 0.14] | 0.001 | 592 |
| | | BMI | 0.75 | -0.02 [-0.11 to 0.06] | < 0.001 | 590 |
| | | BMI *z*-score | 0.49 | -0.03 [-0.11 to 0.05] | 0.001 | 590 |
| **Putamen** | DBSI-FF | WC | 0.55 | -0.03 [-0.11 to 0.06] | 0.001 | 597 |
| | | BMI | 0.21 | 0.05 [-0.03 to 0.14] | 0.003 | 595 |
| | | BMI *z*-score | 0.20 | 0.06 [-0.03 to 0.14] | 0.003 | 595 |
| | DBSI-RF | WC | 0.53 | 0.03 [-0.06 to 0.12] | 0.001 | 595 |
| | | BMI | 0.29 | -0.05 [-0.13 to 0.03] | 0.002 | 593 |
| | | BMI *z*-score | 0.76 | -0.01 [-0.10 to 0.07] | < 0.001 | 593 |

**Supplementary Table 4. Associations between baseline striatal and hypothalamic DBSI metrics and one-year change (Δ) in obesity-related measures.** Standardized (Std.) *β* regression coefficients were reported with 95% confidence intervals (CIs). Analyses were adjusted for baseline obesity-related measures, age at one-year, sex, race/ethnicity, parental education, household income, parental marital status, pubertal development stage at one-year, mean head motion, intracranial volume, and family nested by site. DBSI, diffusion basis spectrum imaging; FF, fiber fraction; RF, restricted fraction; WC, waist circumference; BMI, body mass index.

| Region | Imaging metric (predictor) | 2-year Δobesity-related measure (response) | *p*-value | Std. *β* (95% CI) | Partial $R^2$ | N |
|---|---|---|---|---|---|---|
| **Hypothalamus** | DBSI-FF | WC | 0.17 | -0.06 [-0.14 to 0.02] | 0.003 | 595 |
| | | BMI | 0.55 | 0.03 [-0.06 to 0.11] | 0.001 | 594 |
| | | BMI *z*-score | 0.72 | -0.02 [-0.10 to 0.07] | < 0.001 | 594 |
| | DBSI-RF | WC | 0.035 | 0.09 [0.01 to 0.18] | 0.008 | 594 |
| | | BMI | 0.79 | -0.01 [-0.10 to 0.07] | < 0.001 | 593 |
| | | BMI *z*-score | 0.52 | 0.03 [-0.07 to 0.11] | 0.001 | 593 |
| **Nucleus accumbens** | DBSI-FF | WC | 0.32 | 0.04 [-0.04 to 0.12] | 0.002 | 598 |
| | | BMI | 0.44 | 0.03 [-0.05 to 0.11] | 0.001 | 597 |
| | | BMI *z*-score | 0.64 | 0.02 [-0.06 to 0.10] | < 0.001 | 597 |
| | DBSI-RF | WC | 0.93 | 0.01 [-0.09 to 0.10] | < 0.001 | 597 |
| | | BMI | 0.33 | -0.05 [-014 to 0.04] | 0.002 | 596 |
| | | BMI *z*-score | 0.54 | -0.03 [-0.12 to 0.05] | 0.001 | 596 |
| **Caudate nucleus** | DBSI-FF | WC | 0.21 | 0.05 [-0.03 to 0.14] | 0.003 | 598 |
| | | BMI | 0.27 | 0.05 [-0.03 to 0.13] | 0.002 | 597 |
| | | BMI *z*-score | 0.30 | 0.04 [-0.04 to 0.13] | 0.002 | 597 |
| | DBSI-RF | WC | 0.86 | 0.01 [-0.09 to 0.11] | < 0.001 | 592 |
| | | BMI | 0.10 | -0.08 [-0.17 to 0.01] | 0.005 | 591 |
| | | BMI *z*-score | 0.06 | -0.08 [-0.16 to 0.00] | 0.006 | 591 |
| **Putamen** | DBSI-FF | WC | 0.41 | 0.04 [-0.05 to 0.12] | 0.001 | 597 |
| | | BMI | 0.18 | 0.06 [-0.03 to 0.14] | 0.003 | 596 |
| | | BMI *z*-score | 0.09 | 0.07 [-0.01 to 0.16] | 0.005 | 596 |
| | DBSI-RF | WC | 0.34 | 0.05 [-0.05 to 0.14] | 0.002 | 595 |
| | | BMI | 0.11 | -0.07 [-0.16 to 0.01] | 0.005 | 594 |
| | | BMI *z*-score | 0.22 | -0.05 [-0.14 to 0.03] | 0.005 | 594 |

**Supplementary Table 5. Associations between baseline striatal and hypothalamic DBSI metrics and two-year change (Δ) in obesity-related measures.** Standardized (Std.) *β* regression coefficients were reported with 95% confidence intervals (CIs). Analyses were adjusted for baseline obesity-related measures, age at two-year, sex, race/ethnicity, parental education, household income, parental marital status, pubertal development stage at two-year, mean head motion, intracranial volume, and family nested by site. DBSI, diffusion basis spectrum imaging; FF, fiber fraction; RF, restricted fraction; WC, waist circumference; BMI, body mass index.

| Region | Imaging metric (predictor) | Baseline obesity-related measure (response) | *p*-value | Std. *β* (95% CI) | Partial $R^2$ | N |
|---|---|---|---|---|---|---|
| **Nucleus accumbens** | RSI-RNI | WC | < 0.001 | 0.38 [0.30 to 0.46] | 0.138 | 599 |
| | | BMI | < 0.001 | 0.36 [0.27 to 0.44] | 0.125 | 599 |
| | | BMI *z*-score | < 0.001 | 0.32 [0.24 to 0.40] | 0.101 | 599 |
| **Caudate nucleus** | RSI-RNI | WC | < 0.001 | 0.17 [0.09 to 0.25] | 0.031 | 594 |
| | | BMI | < 0.001 | 0.15 [0.07 to 0.23] | 0.025 | 594 |
| | | BMI *z*-score | 0.001 | 0.13 [0.05 to 0.21] | 0.019 | 594 |
| **Putamen** | RSI-RNI | WC | < 0.001 | 0.18 [0.10 to 0.27] | 0.034 | 600 |
| | | BMI | < 0.001 | 0.17 [0.09 to 0.26] | 0.030 | 600 |
| | | BMI *z*-score | 0.001 | 0.14 [0.06 to 0.22] | 0.020 | 600 |

**Supplementary Table 6. Associations between striatal RSI-RNI and obesity-related measures at baseline.** Standardized (Std.) *β* regression coefficients were reported with 95% confidence intervals (CIs). Analyses were adjusted for age, sex, race/ethnicity, parental education, household income, parental marital status, pubertal development stage, mean head motion, intracranial volume, and family nested by site. RSI, restriction spectrum imaging; RNI, restricted normalized isotropic; WC, waist circumference; BMI, body mass index.

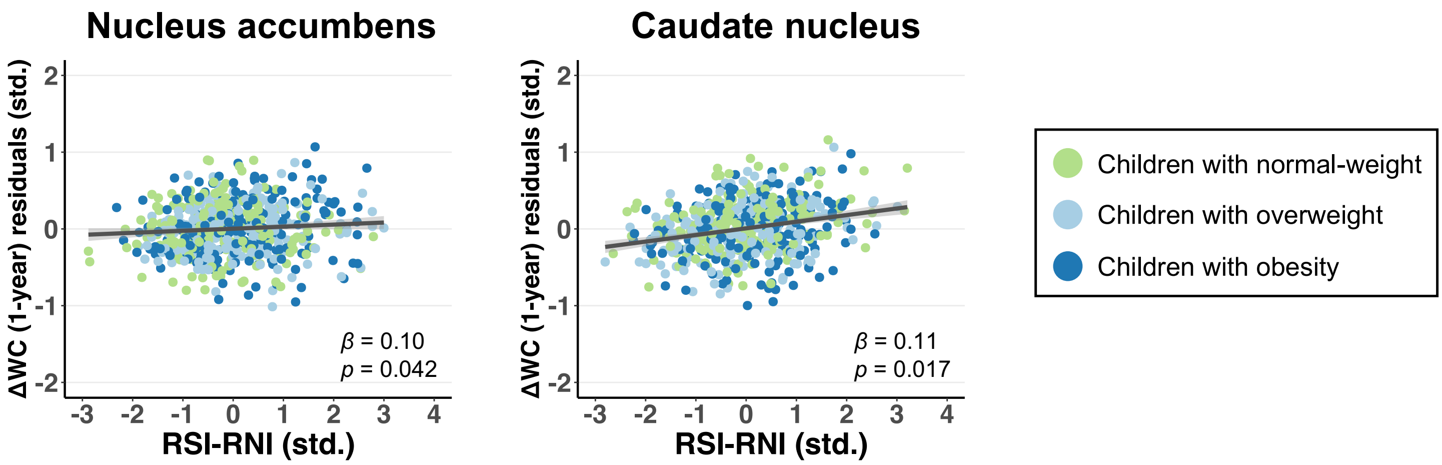


**Supplementary Fig. 5.** Association between baseline RSI-RNI in the nucleus accumbens and caudate nucleus and gain in waist circumference (WC) over one year. WC change (Δ) residuals (adjusted for baseline WC, age at one-year, sex, race/ethnicity, parental education, household income, parental marital status, pubertal development stage at one-year, mean head motion, intracranial volume, and family nested by site) and RSI-RNI were standardized (std.). Standardized $\beta$ regression coefficients were reported with 95% confidence intervals (shaded). RSI, restriction spectrum imaging; RNI, restricted normalized isotropic.

| Region | Imaging metric (predictor) | 1-year Δobesity-related measure (response) | *p*-value | Std. *β* (95% CI) | Partial $R^2$ | N |
|---|---|---|---|---|---|---|
| **Nucleus accumbens** | RSI-RNI | WC | 0.042 | 0.10 [0.00 to 0.20] | 0.008 | 599 |
| | | BMI | 0.22 | 0.06 [-0.05 to 0.16] | 0.003 | 597 |
| | | BMI *z*-score | 0.33 | 0.05 [-0.06 to 0.14] | 0.002 | 597 |
| **Caudate nucleus** | RSI-RNI | WC | 0.017 | 0.11 [0.02 to 0.19] | 0.011 | 594 |
| | | BMI | 0.15 | 0.06 [-0.03 to 0.15] | 0.004 | 592 |
| | | BMI *z*-score | 0.66 | 0.02 [-0.07 to 0.10] | < 0.001 | 592 |
| **Putamen** | RSI-RNI | WC | 0.14 | 0.07 [-0.02 to 0.17] | 0.004 | 600 |
| | | BMI | 0.49 | 0.03 [-0.06 to 0.12] | 0.001 | 598 |
| | | BMI *z*-score | 0.38 | 0.04 [-0.05 to 0.13] | 0.001 | 598 |

**Supplementary Table 7. Associations between baseline striatal RSI-RNI and one-year change (Δ) in obesity-related measures.** Standardized (Std.) *β* regression coefficients were reported with 95% confidence intervals (CIs). Analyses were adjusted for baseline obesity-related measures, age at one-year, sex, race/ethnicity, parental education, household income, parental marital status, pubertal development stage at one-year, mean head motion, intracranial volume, and family nested by site. RSI, restriction spectrum imaging; RNI, restricted normalized isotropic; WC, waist circumference; BMI, body mass index.

| Region | Imaging metric (predictor) | 2-year Δobesity-related measure (response) | *p*-value | Std. *β* (95% CI) | Partial $R^2$ | N |
|---|---|---|---|---|---|---|
| **Nucleus accumbens** | RSI-RNI | WC | 0.10 | 0.08 [-0.02 to 0.18] | 0.005 | 599 |
| | | BMI | 0.30 | 0.05 [-0.05 to 0.15] | 0.002 | 598 |
| | | BMI *z*-score | 0.26 | 0.06 [-0.05 to 0.15] | 0.002 | 598 |
| **Caudate nucleus** | RSI-RNI | WC | 0.24 | 0.05 [-0.03 to 0.14] | 0.003 | 594 |
| | | BMI | 0.36 | 0.04 [-0.05 to 0.13] | 0.001 | 593 |
| | | BMI *z*-score | 0.77 | 0.01 [-0.08 to 0.10] | < 0.001 | 593 |
| **Putamen** | RSI-RNI | WC | 0.47 | 0.04 [-0.06 to 0.13] | 0.001 | 600 |
| | | BMI | 0.81 | 0.01 [-0.08 to 0.10] | < 0.001 | 599 |
| | | BMI *z*-score | 0.89 | -0.01 [-0.10 to 0.08] | < 0.001 | 599 |

**Supplementary Table 8. Associations between baseline striatal RSI-RNI and two-year change (Δ) in obesity-related measures.** Standardized (Std.) *β* regression coefficients were reported with 95% confidence intervals (CIs). Analyses were adjusted for baseline obesity-related measures, age at two-year, sex, race/ethnicity, parental education, household income, parental marital status, pubertal development stage at two-year, mean head motion, intracranial volume, and family nested by site. RSI, restriction spectrum imaging; RNI, restricted normalized isotropic; WC, waist circumference; BMI, body mass index.

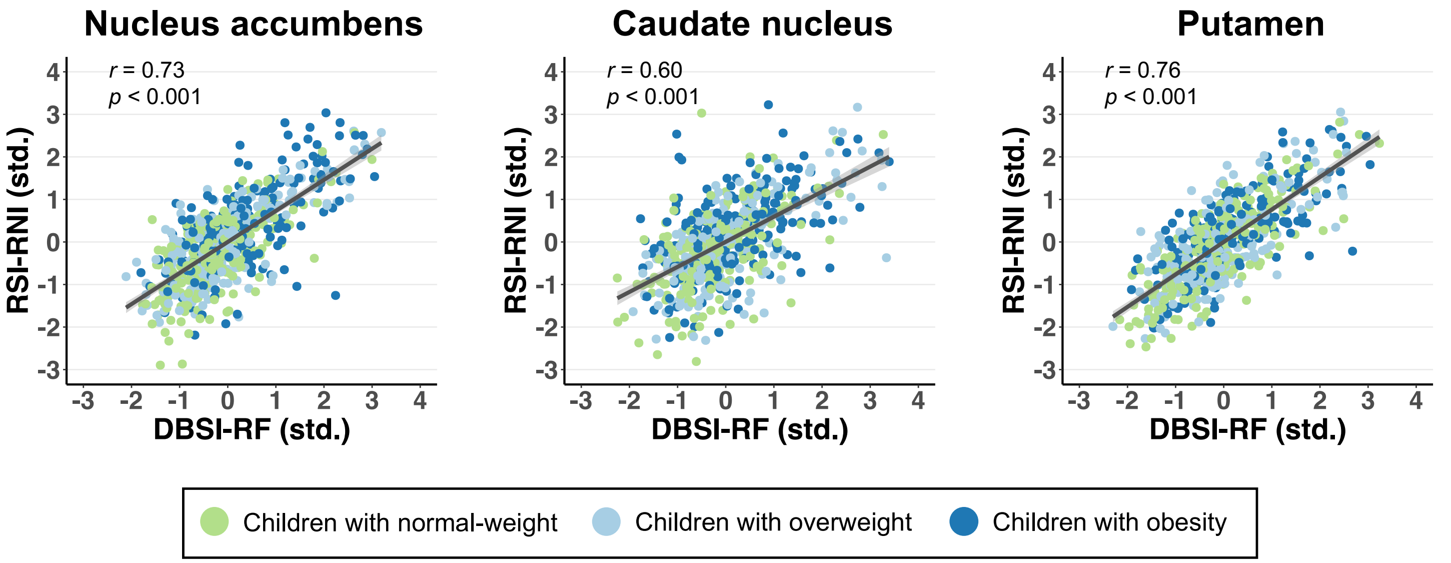


**Supplementary Fig. 6.** Bivariate correlations between DBSI-RF and RSI-RNI in the striatum. Shaded regions represent 95% confidence interval. Correlations were reported as Pearson's *r* coefficients.
